# Longitudinal multi-omic network rewiring at the complement– coagulation interface in post-acute sequelae of COVID-19 (PASC)

**DOI:** 10.64898/2026.07.14.26358048

**Authors:** Bradley Ward, Jean-Luc Balligand, Laurence Bamps, Guido Bommer, Patrice D. Cani, Julien De Greef, Joseph P. Dewulf, Laurent Gatto, Vincent Haufroid, Benoît Kabamba, Sébastien Pyr dit Ruys, Didier Vertommen, Jean Cyr Yombi, Leïla Belkhir, Laure Elens

## Abstract

**Background:** Post-acute sequelae of COVID-19 (PASC) is clinically heterogeneous and mechanistically unresolved, and single-analyte studies have struggled to explain it.

**Methods:** We profiled matched plasma proteomics, metabolomics and whole-blood transcriptomics at acute infection and convalescence (mean 86 days later) in a Belgian cohort, using linear mixed models, multi-omic gene-set enrichment, and a degree-matched differential-correlation approach to quantify how each node’s interactions were rewired between patients who developed PASC and those who recovered; seven axis proteins were additionally quantified by multiplex immunoassay as orthogonal validation.

**Findings:** Single-omic testing yielded few FDR-significant features, yet multi-omic enrichment showed sustained complement-cascade involvement from acute illness to follow-up in PASC. Correlation networks re-organised topologically toward C3 and lost the immunoglobulin V-gene co-expression seen in recovery. The most rewired nodes — heparin cofactor II (SERPIND1), alpha-1 antitrypsin (SERPINA1), complement factor H-related 5 (CFHR5), prothrombin/thrombin (F2) and immunoglobulin V-gene transcripts (notably IGLV3-21) — changed in their co-expression structure rather than in abundance. In multiplex validation, acute CRP was elevated in patients who developed PASC (FDR = 0.012), whereas the directly measured abundances of the network-nominated proteins were unchanged.

**Interpretation:** These trajectory-aware, cross-omic networks nominate a thrombo-inflammatory axis in which complement and coagulation regulation remain dysregulated in PASC at the level of wiring rather than abundance, providing a systems framework for validation and for exploring interventions at the complement–coagulation–platelet interface.

**Funding:** Sofina COVID Solidarity Fund (King Baudouin Foundation), Fondation Saint-Luc and the FNRS.

**Research in context:** *Evidence before this study:* Persistent complement activation, coagulation abnormalities and endothelial dysfunction have been reported in post-acute sequelae of COVID-19 (PASC), and earlier multi-omic studies linked complement and coagulation to persistent symptoms. Most of this evidence, however, rests on differences in the abundance of individual molecules, is cross-sectional or limited in omic breadth, and reports only small effects in convalescent plasma, leaving unclear how these systems are jointly reorganised over time.

*Added value of this study:* Using matched proteomic, metabolomic and transcriptomic profiling of the same plasma at acute infection and convalescence, we quantified how the interaction structure of the complement–coagulation network is rewired between patients who develop PASC and those who recover. This trajectory-aware, network-rewiring readout identified coordinated changes in complement and coagulation regulators (heparin cofactor II, alpha-1 antitrypsin, complement factor H-related 5, prothrombin/thrombin) and immunoglobulin V-gene transcripts that were invisible to single-analyte testing, and a targeted multiplex assay confirmed elevated acute CRP in patients who later developed PASC while showing that the network-nominated proteins did not differ in abundance.

*Implications of all the available evidence:* PASC may be characterised less by large shifts in individual molecules than by a reorganisation of how complement, coagulation and humoral components co-vary. A network-rewiring approach can nominate mechanistically coherent, testable targets at the complement–coagulation–platelet interface and points to the intensity of acute inflammation as a marker of later risk. These hypotheses require confirmation in larger, independent cohorts with orthogonal, quantitative assays before any clinical application.

## Introduction

Coronavirus disease 2019 (COVID-19) is caused by severe acute respiratory syndrome coronavirus 2 (SARS-CoV-2), which enters host cells through interactions with angiotensin-converting enzyme 2 (ACE2) receptors (1). Following infection, SARS-CoV-2 rapidly replicates in respiratory epithelial cells, triggering innate immune activation and inflammatory responses. Clinically, COVID-19 presents across a broad spectrum, ranging from asymptomatic infection to mild flu-like illness, severe pneumonia and multi-organ failure (2).

Furthermore, a significant proportion of patients go on to develop post-acute sequelae of COVID-19 (PASC), commonly referred to as “long COVID,” which manifests itself as persistent fatigue, cognitive impairment, and organ-specific dysfunction months after recovery (3).

Controlled studies estimate PASC prevalence at 8.5–26.4% in adults depending on case definition; hospitalised patients are at markedly higher risk than non-hospitalised, at roughly 54% versus 34% in one global meta-analysis (4,5).

Definitions of PASC vary by study or organization in terms of how long symptoms must persist following acute disease. The World Health Organization (WHO) defines PASC as symptoms at least three months after infection, whilst the UK National Institute for Health and Care Excellence (NICE) and the National Institutes of Health (NIH) RECOVER initiative recognize PASC as early as four weeks post-infection (6–8). Clinical manifestations of PASC are diverse (respiratory, cognitive, cardiovascular, gastrointestinal, etc) and can involve multiple organ systems simultaneously in a relapsing-remitting pattern, reflecting its complexity. (6,9,10).

Multiple, non-mutually-exclusive mechanisms have been proposed for PASC, including viral persistence, autoantibody production, endothelial injury with microthrombosis, and sustained cytokine elevation and immune-cell reprogramming (11–14). Persistent immune dysregulation and chronic inflammation receive substantial support from clinical studies and consortium reviews (e.g. RECOVER) (7,15,16). Recent work also implicates transferable IgG in PASC symptomatology, relevant to the immunoglobulin signal reported here (17–19)

Single-omic studies each capture only part of this multifactorial biology; integrating proteins, metabolites and transcripts from the same patients can reveal convergent pathways. Prior multi-omic studies advanced this, but many had limited integration, lacked longitudinal data, or under-represented non-hospitalised cases (20–24). Rather than re-establishing that complement and coagulation are perturbed in PASC, which has already been shown by abundance-based studies such as Wei et al. (2024) and Cervia-Hasler et al. (2024), we ask how their regulatory interactome is reorganised, using matched tri-omics from a single blood draw, longitudinal acute-to-convalescent sampling, a balanced moderate/severe design, and a degree-matched differential-correlation (rewiring) readout.

## Methods

A full methodological description is provided in Supplementary information 1 for wet- and dry-lab protocols and bioinformatic scripts.

### Patient cohort composition, sample collection, and storage

Participants for this analysis were selected from the HYGIEIA cohort (26): 25 healthy individuals, 24 influenza patients, 50 moderate COVID-19 cases (non-hospitalised, WHO Clinical Progression Scale <4) and 57 severe cases (hospitalised, WHO score ≥4), recruited from Belgian hospitals between August 2020 and March 2024, a window spanning multiple SARS-CoV-2 waves and variants (see Limitations). Samples and clinical data were collected at inclusion and again at convalescent follow-up (mean 86.2 days after inclusion, SD 30.5, range 60–274). The study was conducted in accordance with the Declaration of Helsinki and approved by the Institutional Review Board of Cliniques Universitaires Saint-Luc (protocol 2021/30DEC/543); it is registered on ClinicalTrials.gov (NCT05557539) and all participants gave written informed consent.

PASC was defined as the presence or continuation of symptoms at visit 2 without an obvious alternative explanation, adjudicated by a study nurse; recovered patients were those without such symptoms at visit 2. Full vaccination was defined as receipt of the recommended vaccine doses prior to inclusion (Janssen, Pfizer–BioNTech, Moderna or AstraZeneca).

### Wet-lab methodology

#### DNA extraction and whole exome sequencing

DNA was extracted from EDTA blood using QIAamp DNA Blood Mini Kit (Qiagen, Germany). WES was outsourced to Macrogen (Republic of Korea) who used the Twist Human Core Exome (+RefSeq) system (Twist Bioscience, USA), sequencing was carried out on a Novaseq 6000 platform to a target depth of 5Gb, 50x mappable, 151 paired end reads.

#### Whole Transcriptomic Shotgun RNAseq

Total RNA was extracted from whole-blood samples using the Tempus™ Spin RNA Isolation Kit (Thermo Fisher Scientific, USA) and purified using the RNA Clean & Concentrator-5 Kit (Zymo Research, USA). RNA-seq was outsourced to Macrogen who used the Illumina Globin-Zero Gold Kit (Illumina, USA), followed by the TruSeq Stranded mRNA protocol (Illumina, USA) for library construction. Normalized pooled libraries were sequenced on a Novaseq 6000 (Illumina, USA) to a target depth of 30M 151 paired end reads.

#### DIA Shotgun Proteomics

Plasma proteomics used data-independent acquisition on an Orbitrap Exploris 240 (Thermo Fisher Scientific) following our published protocol (27). Briefly, Plasma samples were depleted via High Select Top14 Abundant Protein Depletion Resin (Thermo Fisher Scientific, USA), and were then denatured, reduced, alkylated, and digested overnight with trypsin (V5117, Promega, United States). Peptides were then fractionated (Pierce High pH Reversed-Phase Peptide Fractionation column, Thermo Fisher Scientific, USA) and eluted into seven fractions which were combined into three distinct pools. 1 µg of peptide was injected into a reversed-phase pre-column (Acclaim PepMap 100, Thermo Fisher Scientific, USA) and separated using an EasySpray analytical column (Acclaim PepMap RSLC C18, 0.075 × 250 mm, Thermo Fisher Scientific, USA) over a 120-minute gradient at 300 nL/min on an Ultimate 3000 RSLC nanoHPLC system (Thermo Fisher Scientific, United States). A data-independent acquisition (DIA) approach was employed at an m/z range of 500–740 with a 4 m/z isolation window at a resolution of 60,000 (FWHM), acquired on an Orbitrap Exploris 240 mass spectrometer (Thermo Fisher Scientific, USA).

#### Shotgun Metabolomics

Metabolomics analysis was performed as previously described (28) on plasma samples following acetonitrile protein precipitation. Metabolites were extracted twice, pooled, dried under nitrogen, and reconstituted for four analytical methods: reverse-phase (HSS T3 column, Waters, USA) and HILIC (BEH Amide column, Waters, USA), each in positive and negative electrospray ionisation. Samples were analysed using an ACQUITY Premier UPLC system coupled to a Synapt-XS high-resolution Q-TOF mass spectrometer (Waters), acquiring data in MSE mode over an m/z range of 50–1200 with leucine-enkephalin lock-mass.

### Bioinformatic preprocessing pipelines

Proteomic spectra were initially processed using DIA-NN (v1.8.1) (29). Missing precursors were filtered or flagged MNAR/MAR and imputed (QRILC/random forest) following recommended practice (30), normalised and summarised to protein level.

Metabolomic data were imported into Progenesis QI (V4.2, Nonlinear Dynamics, UK) for lock mass correction and 2D ion intensity map conversion. Following feature detection, identification, and alignment, raw abundance values were then filtered and imputed as previously described for proteomics, normalized, and batch corrected.

Transcriptomic raw fastq files were first trimmed to remove low-quality bases (Trimmomatic v0.39) and reads were aligned to the GRCh38 human genome (HISAT2 v2.2.1) (31,32). Gene-level expression was quantified with featureCounts (Subread v2.0.3) using the GRCh38.105 genome assembly (33). Normalized count matrices were generated using DESeq2 (34).

Genomic reads were aligned to GRCh38 human genome (BWA-mem v0.7.17) (35), followed by duplicate removal (SAMTools v1.12) and base quality recalibration (GATK BaseRecalibrator v4.2.1) (36,37). Germline mutations were called using GATK HaplotypeCaller (37). Structural variations were detected by combining “read depth” (ExomeDepth) with “split read” (GRIDSS-Purple-Linx) (38,39). Resulting files were normalized, filtered, and annotated with snpEff (v5.0e) (37,40).

### Analytical methodologies

Genomic SNP data was filtered according to strict QC metrics, per-sample and SNP call rates, and predictive damaging scores (as calculated within Highlander); remaining missing genotypes were mean-imputed per SNP. PCA was computed on LD-pruned, position-sorted genotypes and PC1 accounted for the most captured variation (2.4%), including ethnicity. For the final GWAS model, we tested the association between SNP dosage (0-2, additive) and PASC status (PASC vs recovered) using logistic regression, adjusting for age (years), sex (male/female), and ancestry (PC1). Following, we tested per-gene burden/variance effects using SKAT-O, run using predictive damaging scores of the SNP.

Proteomic and metabolomic abundances and RNA-seq counts were modelled with linear mixed models (dream/variancePartition), testing the PASC×Visit interaction with a random intercept for patient and covariates (severity, vaccination, sex, age); cross-sectional contrasts used limma. Multi-omic pathway enrichment used multiGSEA across KEGG and Reactome: for each omic, features were ranked by a signed significance score (direction of the PASC/Visit effect × -log10 p) and tested against KEGG and Reactome gene- and metabolite-sets by per-omic gene-set enrichment (fgsea); per-pathway enrichment p-values were then aggregated across the three omics by the Z-method (Stouffer’s) and Benjamini–Hochberg-adjusted across all tested pathways, with pathways at combined FDR < 0.05 considered significant.

MultiGSEA results highlighted specific Reactome pathways “Complement Cascade” and the sub-pathway “Regulation of Complement Cascade”. All proteins, transcripts, and metabolites for this network were selected as seed nodes and the network was then expanded to included first-neighbours of seed nodes from a curated “prior knowledge” network.

For the complement-cascade-seeded network, per-omic differential correlations between PASC and recovered were computed (DGCA), and a degree-matched differential-correlation (rewiring) score was derived per node by combining its edge changes (L2 norm) and z-scoring within degree-matched bins. Because the degree-matched z-score (DMZ) compares each node only against peers of similar degree, a high score reflects coordinated rewiring rather than the number of connections a node has or the accumulation of many negligible, random edge changes.

Networks were built for the visit-2 contrast (n = 32 PASC, 45 recovered); the cross-sectional contrasts had 45 recovered and 29 PASC at visit 1, and 34 recovered and 17 PASC at visit 2.

### Targeted multiplex validation

As an orthogonal, quantitative validation of the complement–coagulation–inflammatory axis nominated by the discovery omics, seven plasma analytes (C9, C-reactive protein (CRP), coagulation factor VII, alpha-1 antitrypsin (SERPINA1), von Willebrand factor, sICAM-1 and sVCAM-1) were quantified by electrochemiluminescence multiplex immunoassay (Meso Scale Discovery) in the same acute (visit 1) and convalescent (visit 2) plasma samples, each measured in duplicate. PASC and recovered patients were compared per analyte at each visit using the Mann–Whitney U test (with a log-scale Welch t-test as sensitivity analysis), and effects were re-estimated in covariate-adjusted linear (log10 concentration) and logistic (PASC status) models using the discovery covariate set (acute severity at visit 1, vaccination, sex and age). The acute response was additionally stratified by severity, and matched patients were analysed with a Visit × PASC linear mixed model mirroring the dream framework.

## Results

### Cohort characteristics

Table 1 summarises the cohort. PASC and recovered patients were broadly similar in age and sex, but PASC was strongly associated with acute severity (71% of PASC patients versus 29% of recovered patients had severe disease), which is therefore included as a covariate in all models.

**Table 1.** Cohort characteristics at inclusion, by convalescent PASC status.

| Characteristic | All (n=76) | Recovered (n=45) | PASC (n=31) |
| --- | --- | --- | --- |
| Age, years — mean (SD) | 50.5 (15.7) | 48.4 (14.9) | 53.5 (16.6) |
| Female sex — n (%) | 37 (49) | 24 (53) | 13 (42) |
| BMI, kg/m<sup>2</sup> — mean (SD) | 25.8 (6.8) | 23.9 (5.2) | 28.7 (7.9) |
| Moderate disease — n (%) | 39 (51) | 31 (69) | 8 (26) |
| Severe disease — n (%) | 35 (46) | 13 (29) | 22 (71) |
| Fully vaccinated — n (%) | 69 (91) | 43 (96) | 26 (84) |
| Dexamethasone — n (%) | 36 (47) | 15 (33) | 21 (68) |
| ICU admission — n (%) | 4 (5) | 1 (2) | 3 (10) |
| Deaths — n | 0 | 0 | 0 |

### Differential expression and GSEA analysis

Using single-omic models adjusted for age, sex, vaccination, and acute severity, no individual features reached FDR < 0.05 across acute (visit 1), convalescent (visit 2), or longitudinal contrasts. Multi-omic GSEA detected coordinated pathway shifts, four of these pathways were significant (combined cross-omic FDR < 0.05) at both visit 1 and visit 2 but not longitudinally, suggesting sustained elevation or depletion in PASC patients vs recovered patients: Complement cascade, Regulation of complement cascade, Intraflagellar transport, and Regulation of tubulin folding by CCT/TriC. Of these pathways, the complement cascade was selected for deeper investigation due to its previously suggested involvement in PASC pathogenesis. Notably, although no single feature reached FDR at any contrast, the two complement pathways survived Benjamini–Hochberg correction across all tested pathways and did so independently at both visits. In the two complement pathways (Fig. 1), proteomic signal drove the enrichment at both timepoints (transcriptomic signal weak; no complement-mapped metabolites).

**Figure 1:**
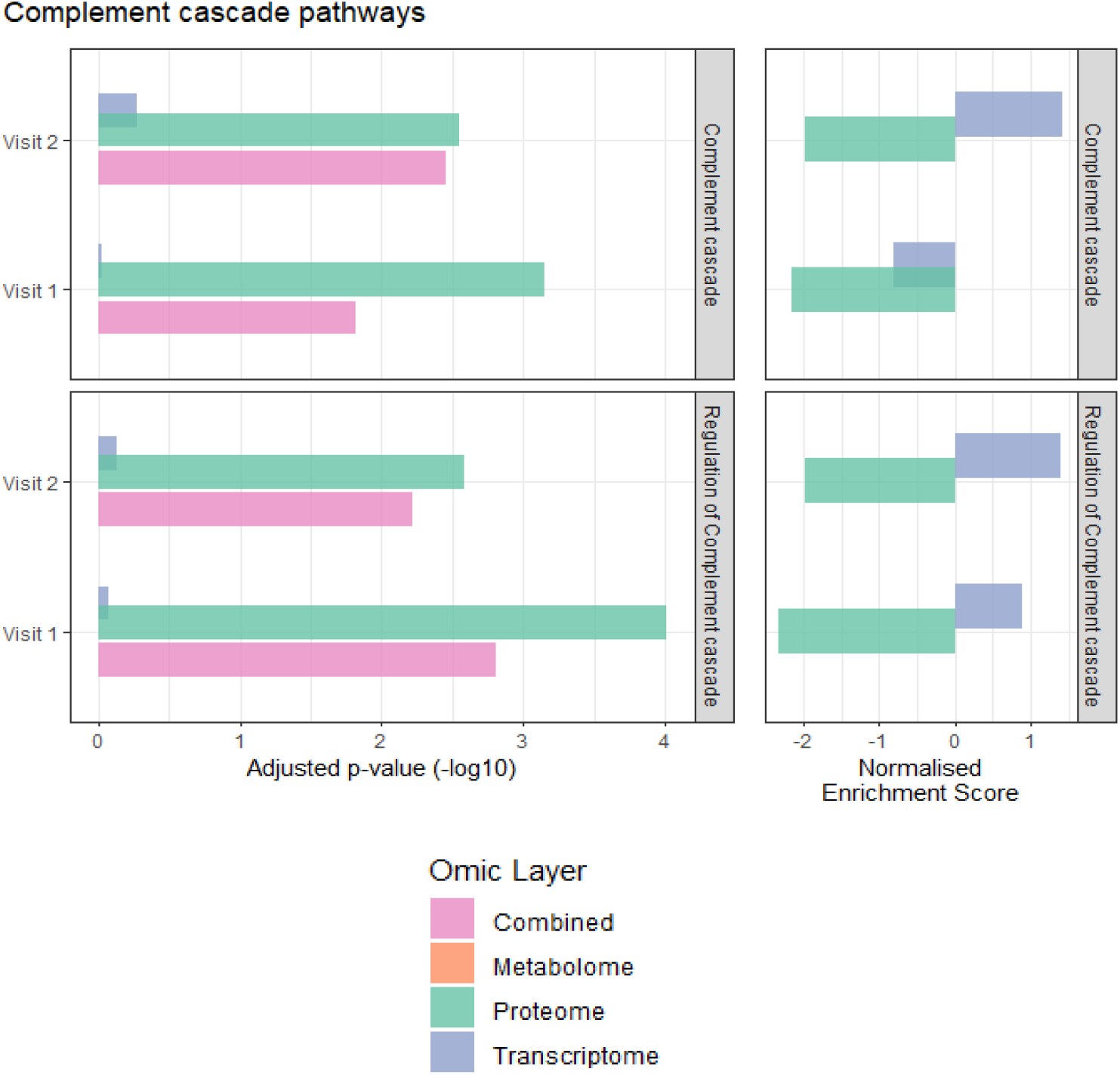
multiGSEA of selected Reactome pathways **Complement Cascade** and **Regulation of Complement Cascade**.

Genome-wide analysis identified no variant reaching genome-wide significance (top SNPs rs767558704/ZNF233, rs17283/TRBV13 and rs1730409324/MUC4; all p ≈ 0.003–0.004, far from the 5 × 10⁻⁸ threshold), and gene-level aggregation (SKAT-O) yielded no gene at FDR < 0.05 (top genes ITPR2, GANC and AC011005.1). No single locus explained PASC status, consistent with the omics and reinforcing a multifactorial, network-level signal.

### Network modules and node reorganisation

We integrated cross-omic interactions pertaining to transcription, translation, transcription factor-gene targets, miRNA-mRNA targets, curated protein-protein and protein-metabolite interactions, and significant within-omic correlations to build visit 2 networks for PASC and recovered groups (Fig. 2A–B respectively). In the recovered network (Fig. 2B), nodes partitioned into two modules: a complement/coagulation module and an immunoglobulin V-gene transcript module. This layout was retained in Figs 2A and 3 to aid visual comparison.

**Figure 2:**
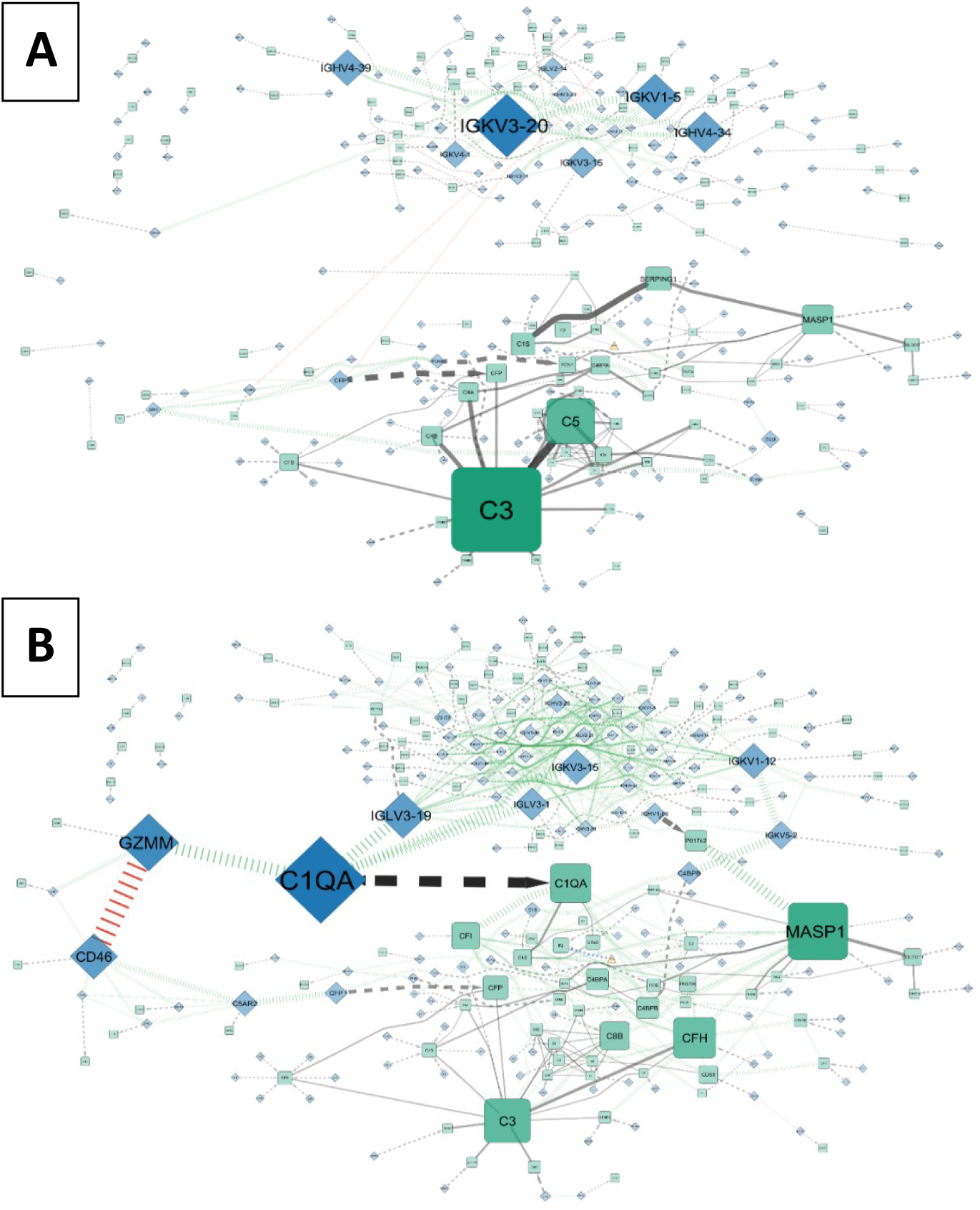
Complement cascade network in PASC patients (A) and recovered patients (B) during convalescent phase of COVID-19. Nodes: Green squares = proteins, blue diamonds = RNA, orange triangles = metabolites. Edges: dashed black edge = transcription, solid black edge = protein-protein interaction, green vertical dashed edge = significant positive correlation, red vertical dashed edge = significant negative correlation, blue dotted edge = protein-metabolite interaction. Larger node size and edge thickness correspond to a higher betweenness score of the node/edge. Node positions were calculated based on prefuse force directed layout algorithm on the recovered network, and then mapped to the PASC network. Network A contains 165 RNA, 113 proteins, and 1 metabolite. Network B contains 165 RNA, 114 proteins, and 1 metabolite.

**Figure 3:**
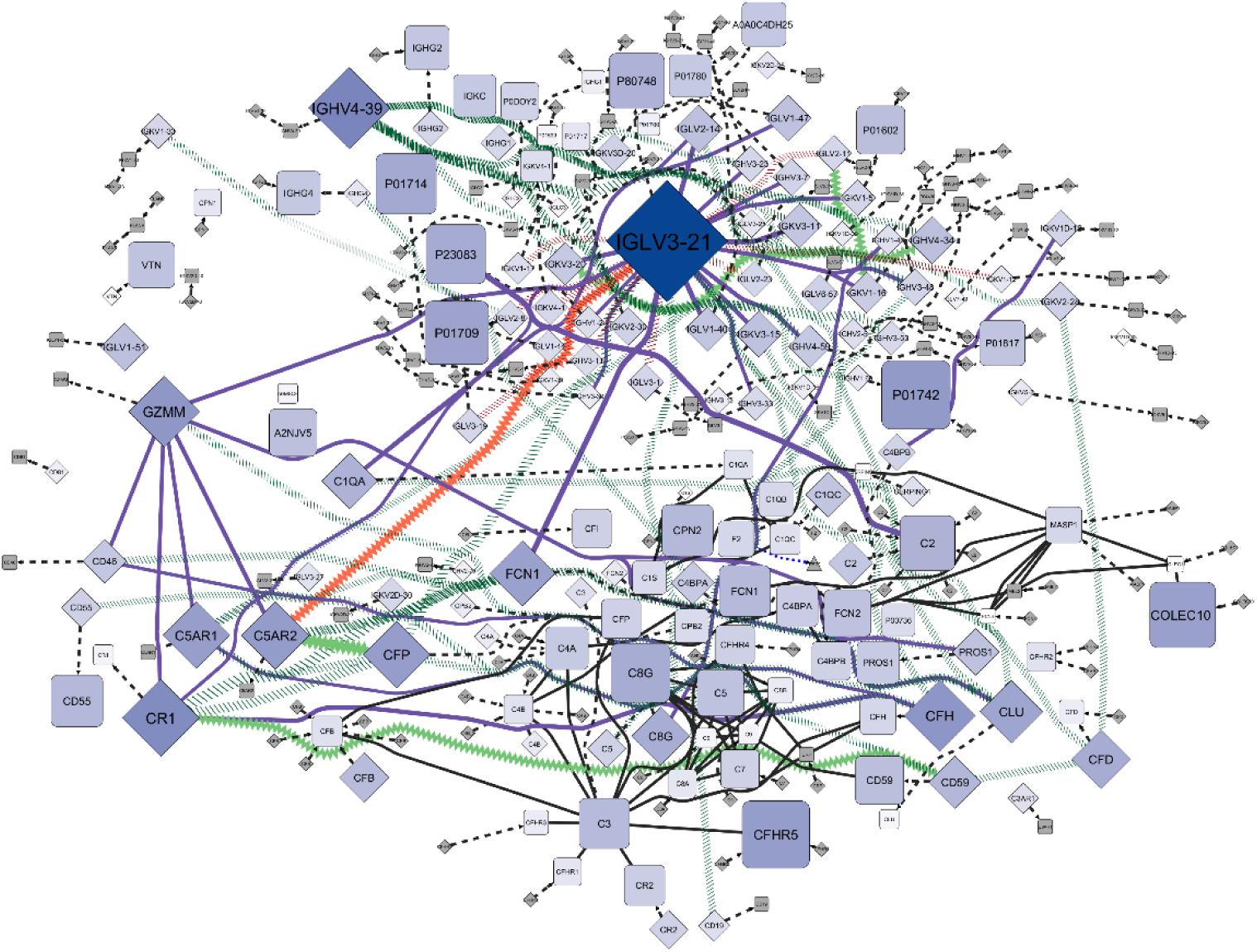
Differential network analysis of the complement cascade network, PASC vs recovered. Nodes: squares = proteins, diamonds = RNA, triangles = metabolites. Edges: dashed black edge = transcription, solid black edge = protein-protein interaction, green vertical dashed edge = correlation gained in PASC, red vertical dashed edge = correlation lost in PASC, green zigzag edge = strengthened positive correlation in PASC, red zigzag edge = strengthened negative correlation in PASC, red sinewave edge = weakened negative correlation in PASC, green sinewave edge = weakened positive correlation in PASC, purple solid edge = correlation sign flip in PASC, blue dotted edge = protein-metabolite interaction. Larger node size and deeper color corresponds to normalized rewiring score, edge thickness corresponds to z-score of correlation difference PASC vs recovered. Node positions were calculated based on prefuse force directed layout algorithm on the recovered network (Fig. 2B). Edges were pruned to only show significant, strong changes (adjusted p-value < 0.05). Network contains 165 RNA, 116 proteins, and 1 metabolite.

Relative to recovery, the PASC network (Fig. 2A) showed a topological re-centring of the complement module toward C3, with betweenness shifting away from the regulatory hubs (MASP1, SERPING1) that were central in recovery; we describe this as a change in network topology rather than molecular flux, and reserve mechanistic interpretation for the Discussion. The recovered immunoglobulin module was densely connected through strong positive co-expression, whereas in PASC most of these transcript correlations were lost, giving a sparser module.

### Differential wiring (PASC vs recovered)

Combining the networks and analysing nodes through the DMZ, immunoglobulin V-gene transcripts accounted for most reorganisation in PASC, with IGLV3-21 the top node (DMZ = 5.33) and IGHV4-39 secondary (DMZ = 2.74) (Fig. 3). The dominant pattern is that IGLV3-21 switches from loosely positive embedded within the IgV cluster in recovered to becoming a negatively correlated hub in PASC. This is a change in co-expression structure, not abundance (IGLV3-21 transcript log-fold-change non-significant), and may connect restricted immunoglobulin-V usage to the autoantibody hypothesis of PASC. Additional nodes that surface through cumulative coordinated edge changes include P01742 (IGHV1-69 product, DMZ = 1.91), CFHR5 (C3 regulator in the alternative pathway, DMZ = 1.84), and COLEC10 (lectin pathway, DMZ = 1.72).

### Regulation within the complement control layer

Within the complement-control layer (figure 3), complement factor H (CFH; DMZ = 2.08) and CFHR5 (DMZ = 1.84) were among the most reorganised regulators, and CR1 the most reorganised node of this group (DMZ = 2.45). We report each node by its rewiring score and the classes of partner it re-couples to (for CFH, other complement regulators and receptors; for CFHR5, many small changes across the alternative-pathway neighbourhood) rather than enumerating individual edges.

#### Broader coagulation and anticoagulant neighbourhoods

In figure 4, first neighbours of the original seed network (figure 3) are included in the analysis. In this broader coagulation neighbourhood, IGLV3-21 still remains the largest rewired node despite no significant abundance change (DMZ = 3.48). Most IGLV3-21 node partners are other immunoglobulin V genes, so the net effect is that IGLV3-21 switches from being loosely/positively embedded within the IgV cluster in recovered to acting as a negatively correlated hub in PASC, including several very strong de-novo negatives.

**Figure 4:**
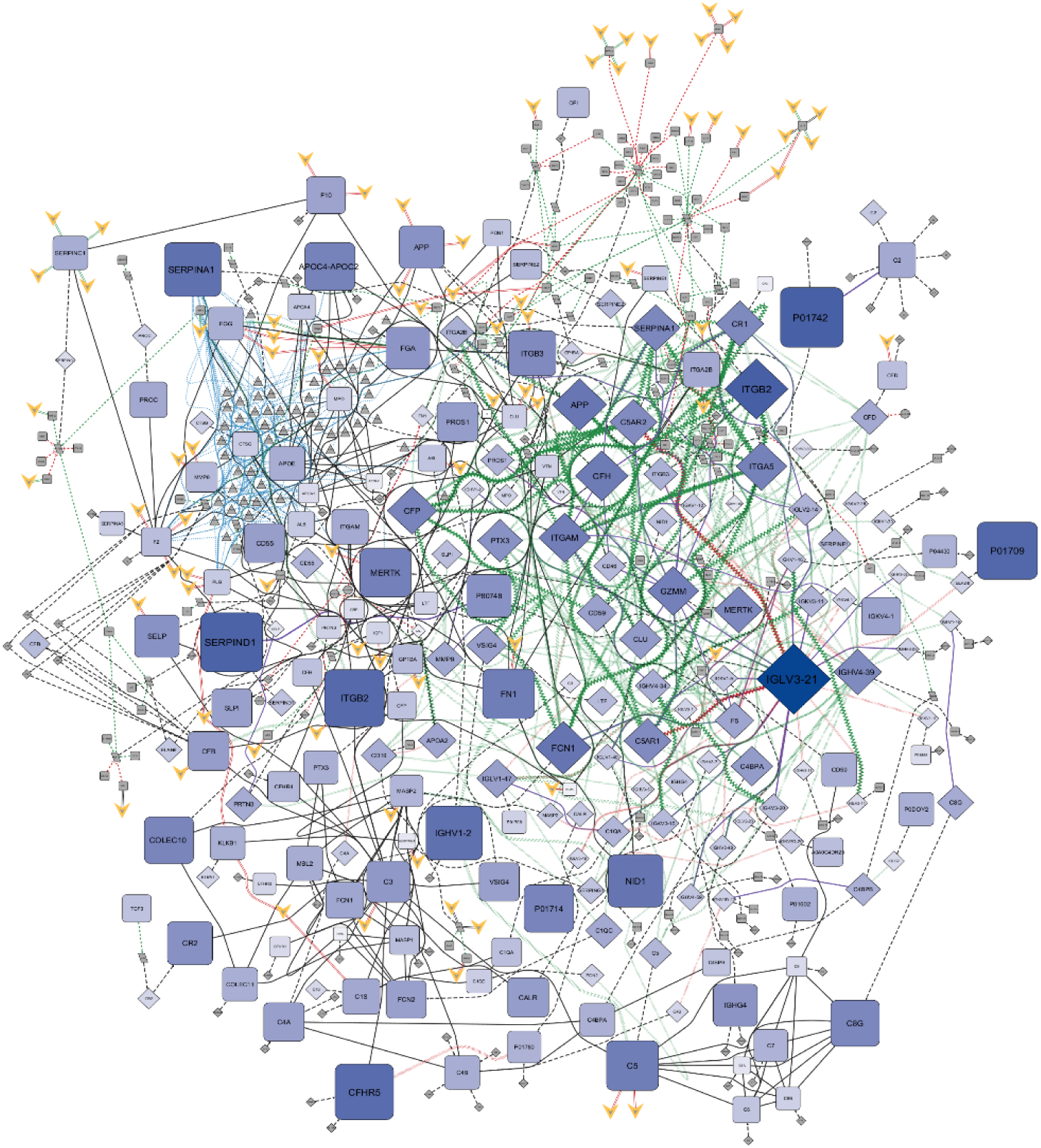
Differential network analysis of the complement cascade network, PASC vs recovered, with non-correlation edge first neighbors. Nodes: squares = proteins, diamonds = RNA, triangles = metabolites, ellipse = miRNA, parallelogram = gene, yellow V = drug. Edges: dashed black edge = transcription, solid black edge = protein-protein interaction, green vertical dashed edge = correlation gained in PASC, red vertical dashed edge = correlation lost in PASC, green zigzag edge = strengthened positive correlation in PASC, red zigzag edge = strengthened negative correlation in PASC, red sinewave edge = weakened negative correlation in PASC, green sinewave edge = weakened positive correlation in PASC, purple solid edge = correlation sign flip in PASC, blue dotted edge = protein-metabolite interaction, green dashed edge = activation, red dashed edge = repression, green parallel edge = drug activation/agonist, red parallel edge = drug repression/antagonist. Larger node size and deeper color corresponds to normalized rewiring score, edge thickness corresponds to z-score of correlation difference PASC vs recovered. Node positions were calculated based on prefuse force directed layout algorithm on the recovered network (Fig. 2B). Edges were pruned to only show significant, strong changes (adjusted p-value < 0.05). Network contain 175 RNA, 5 miRNA, 205 proteins, 49 metabolites, 32 genes, and 77 drugs.

Together, these patterns appear to point to a reorganization of IgV co-expression structure around IGLV3-21 rather than a simple up- or downregulation of the transcript itself. Other changes of note within the regulatory environment include heparin cofactor II (SERPIND1; protein DMZ = 2.27), alpha-1 antitrypsin (SERPINA1; RNA DMZ = 1.33) and integrin β2 (ITGB2/CD18; DMZ = 2.27 protein, 2.58 RNA), which were highly reorganised, and prothrombin (F2), which itself showed little reorganisation (DMZ = −0.27) and no significant abundance change; the reorganisation instead concentrates in its regulators. Anticoagulants (SERPINC1, PROC) also trended lower without reaching significance. We place individual node rewiring and edge statistics in Supplement table 1 and 2 respectfully.

CRP shows a trend to higher protein (logFC = 0.98, non-adjusted p-value = 0.13) but very low rewiring (DMZ = −1.10), and the transcript is likewise stable and only linked to its protein. Within the multi-omic network, CRP forms only three physical edges without any significant or strong correlations present in either PASC or recovered networks, indicating that the manner in which CRP interacts with the wider complement and coagulation network is essentially unchanged in PASC patients. This does not contradict the multiplex validation reported below: abundance-based assays and the rewiring score capture different phenomena, and CRP’s acute abundance signal need not manifest as a change in network wiring.

#### Targeted multiplex validation of the complement–coagulation axis

Of seven analytes, only CRP differed significantly between groups: at acute infection CRP was ∼4.5-fold higher in patients who subsequently developed PASC (Mann–Whitney p = 0.0017, FDR = 0.012), the only analyte to survive correction; the direction persisted after adjustment (adjusted OR ≈ 1.9 per SD of log10-CRP, p ≈ 0.05) and attenuated to borderline when conditioning on acute severity (itself strongly associated with both CRP and PASC). The signal localised to moderately ill patients (moderate-PASC CRP ∼5-fold above moderate-recovered, adj. p = 0.010; indistinguishable from severe patients, adj. p = 0.41), whereas severe patients had uniformly high CRP irrespective of outcome (adj. p = 0.34).

The remaining six analytes, including the network-nominated SERPINA1 and C9, showed no significant abundance difference between PASC and recovered patients at either visit, although CRP and factor VII showed nonsignificant upward trends in PASC at convalescence (adjusted p ≈ 0.06–0.08). In the matched longitudinal model, acute CRP fell sharply between visits in both groups with no significant Visit × PASC interaction (adj. p ≈ 0.19), indicating comparable resolution; the only analyte trending toward differential non-resolution was sICAM-1, which declined in recovered patients but remained flat in PASC (interaction adj. p ≈ 0.12) (Fig. 5; full per-analyte results in Supplementary Table 3).

**Figure 5.**
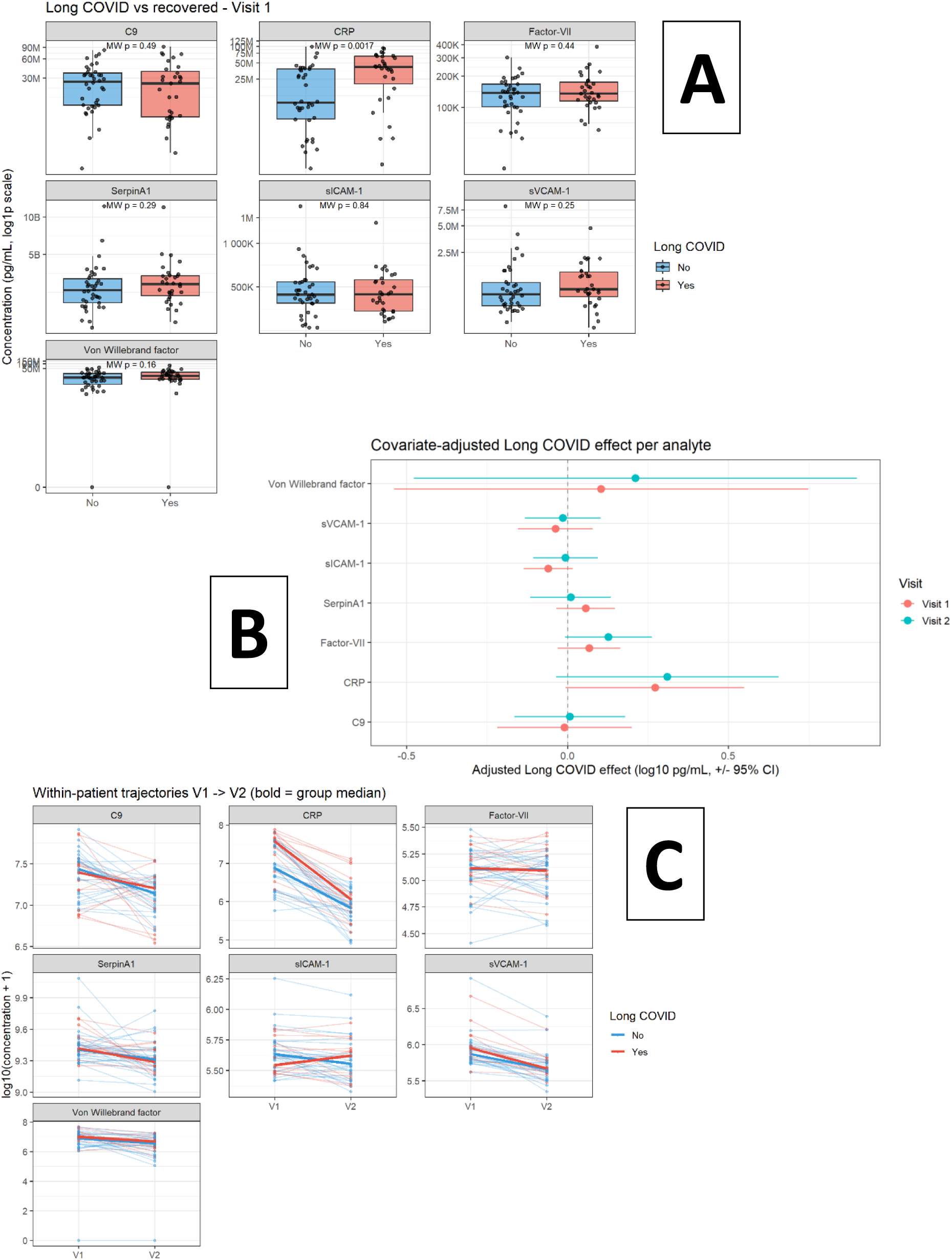
Targeted multiplex (Meso Scale Discovery) validation of seven plasma analytes. (A) Acute (visit 1) CRP in PASC versus recovered patients across all COVID-19 cases (box-and-jitter with the Mann–Whitney p-value); (B) the same, stratified by acute severity and PASC status, showing moderate-PASC patients overlapping the severe range and recovered-moderate patients well below; (C) matched within-patient CRP trajectory from visit 1 to visit 2 by group.

## Discussion

This exploratory multi-omic analysis nominates the complement–coagulation interface as a candidate system for targeted follow-up in PASC, on three convergent signals: pathway-level enrichment at both acute and convalescent phases; node-level reorganisation of complement and coagulation regulators; and orthogonal confirmation of an acute inflammatory (CRP) signal, despite limited differential abundance at the single-feature level. As shown by our network analysis (Fig. 4), key coagulation and complement components exhibit extensive rewiring in PASC. Figure 6 has been created in an effort to summarise and explain the main findings of our investigations, showing how some of the nodes we have identified as most rewired or interesting (highlighted in yellow) fit into the complex and highly interconnected coagulation and complement pathways.

**Figure 6:**
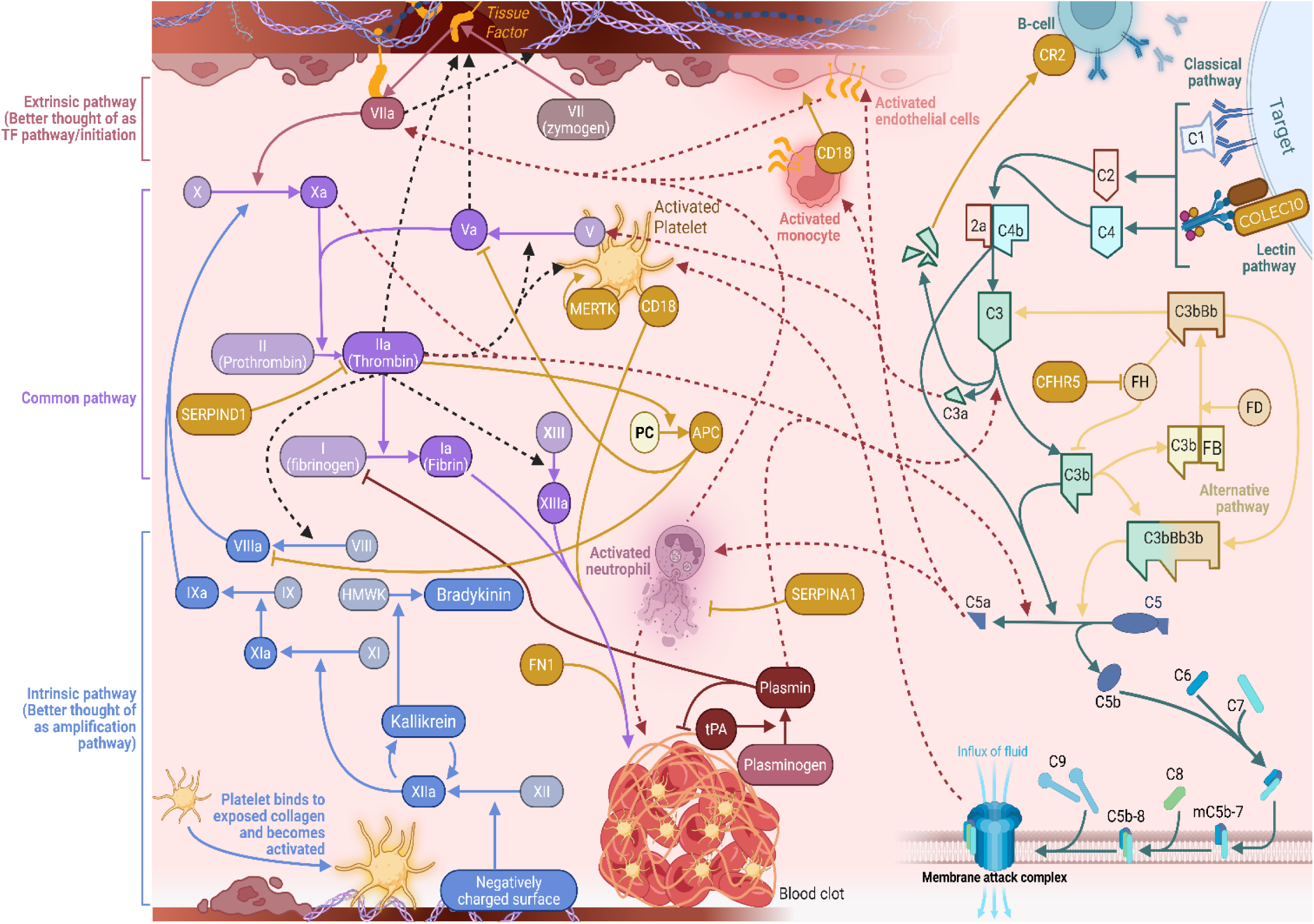
Complement coagulation pathways with interactions and proteins of interest. **APC:** When thrombin binds to thrombomodulin on intact endothelium, it activates protein C (APC) which inactivates Va and VIIIa, shutting down thrombin generation. Losing this pathway tilts you towards a procoagulant environment. **SERPIND1.** Heparin cofactor II (SERPIND1) inhibits thrombin (IIa). Loss of this inhibition increases thrombin activity. **SERPINA1:** α1-antitrypsin (SERPINA1) inhibits neutrophil elastase (released during NETosis), limiting neutrophil-mediated tissue damage. **MerTK:** Platelets express TAM (Tyro3, Axl, and Mer) receptors like MerTK. MerTK amplifies platelet activation upon binding with GAS6 (released by endothelial cells and macrophages after stimulation by factors such as thrombin, collagen, etc). **FN1:** Plasma fibronectin (FN1) gets crosslinked into fibrin clots by factor XIIIa which improves clot stability, platelet adhesion to the fibrin network, and supports wound repair. **CD18/ITGB2:** CD18/Integrin β2 (ITGB2) pairs with α subunits (like ITGAM/CD11b) to form Mac-1, which on activated neutrophils/monocytes binds fibrinogen/fibrin and iC3b (C3b breakdown product),and also binds ICAM-1 on activated endothelium. This lets leukocytes dock onto thrombi and onto inflamed endothelium. **CFHR5:** Complement Factor H-Related protein 5 (CFHR5) regulates the complement system by modulating factor H’s inhibitory activity against C3b and other binding targets, attenuating its effect on complement activation. Dysregulation here could increase or decrease the amplification step of complement activation. **CR2**: Complement receptor 2/CD21 (CR2) on B cells binds C3 breakdown products (C3d/iC3b/C3dg) which lowers the threshold for B cell activation and helps retain immune complexes, feeding antibody production, which then feeds the classical pathway via C1q binding IgG/IgM. Dysregulation here could affect antibody production and classical complement activation. **COLEC10:** Collectin-10 (COLEC10), also called collectin liver-1 or CL-L1, is a soluble pattern-recognition lectin in the lectin pathway that forms complexes with MASPs. It binds mainly with mannose, fucose, and galactose. Dysregulation here could result in abhorrent lectin pathway activation. Created in BioRender. Elens, L. (2025) https://BioRender.com/w1ylbg9

The picture is of a reorganised thrombo-inflammatory interface rather than up- or down-regulation of individual factors. Highly reorganised nodes span humoral immunity (IGLV3-21), complement regulation (CFH, CFHR5), coagulation control (SERPIND1, SERPINA1, PROC), leukocyte–platelet adhesion (ITGB2/CD18) and matrix/endothelial integrity (FN1, NID1). We reiterate that we interpret these patterns as hypotheses: the data are correlation-based, with non-significant abundance changes, and cannot establish thrombin generation, platelet activation or endothelial injury directly; each mechanistic step is inferred, not measured.

Analysis of the rewiring network, considering the biological context, shows that the reorganisation concentrates in thrombin’s regulatory environment rather than in prothrombin (F2) itself. Notably, several natural anticoagulants that restrain thrombin were downregulated (non-significantly) or dysregulated across omics: heparin cofactor II (SERPIND1) emerged as one of the most rewired proteins in the entire network, while antithrombin (SERPINC1) and protein C (PROC) trended lower at both RNA and protein levels. SERPIND1’s role is to neutralize thrombin and apply a negative regulation to coagulation activity (41). Dysregulation could theoretically lead to increased thrombin on healthy surfaces, favouring aberrant clot formation and increased C5 and C3 cleavage. C3 and C5 fragments then lead to activation of monocytes, neutrophils, endothelial cells, and platelets, which further activates complement and coagulation pathways as seen in figure 6 (42). PROC should curb this thrombin generation through inactivation of Va, which prevents thrombin activation directly, and through inactivation of VIIIa, which shuts down the amplification/intrinsic pathway of thrombin generation (43). On the other hand, CFHR5 competes with factor H (an inhibitor of the alternative complement pathway), thereby de-repressing complement activation (44). C3 fragments also go on to bind to CR2 on B cells, lowering their threshold for activation and increasing antibody production, yet a large number of antibody components also showed significant rewiring (45).

Indeed, IGLV3-21 was the most rewired/dysregulated node and may imply dysregulation, compared to recovered patients, in the B-cell/antibody activity and the thrombin-coagulation axis in PASC. Consistently, we observed that integrin β2 (ITGB2, CD18), a leukocyte adhesion molecule and component of complement receptor 3, became highly rewired in PASC. CD18 is involved in allowing leukocytes to dock onto thrombi or inflamed endothelium (46). Its new network orientation indicates enhanced coordination between leukocyte integrin signalling, complement, and coagulation pathways, potentially facilitating platelet-leukocyte aggregate formation and immune-thrombotic feedback loops unique to PASC when compared to recovered patients. Furthermore, markers of endothelial and matrix integrity reflected ongoing vascular perturbation: fibronectin (FN1), a high-molecular-weight glycoprotein that stabilises fibrin clots, and nidogen-1 (NID1), a basement membrane protein, were modestly rewired in PASC.

The multiplex immunoassay provides orthogonal support for the inflammatory component: whereas discovery proteomics detected only a non-significant CRP trend at visit 1 (log-fold-change 0.98, unadjusted p = 0.13), the immunoassay confirms an FDR-significant elevation of acute CRP in patients who later develop PASC. Because PASC status is defined at follow-up, this is best read as a predictive marker of acute inflammatory intensity that largely resolves by convalescence — concordant with the low convalescent rewiring of CRP in our network. The absence of an abundance difference for SERPINA1, C9 and the coagulation/endothelial markers is consistent with (though not proof of) a model in which these components are altered in their correlation structure rather than their mean levels; testing this will require protein-complex or activation-state assays (e.g. CFHR5/SERPIND1 ELISA, complement activation fragments), as CFHR5 and SERPIND1 were not on the validation panel.

Taken together, these findings suggest that PASC is a state of persistent thrombo-inflammatory disequilibrium: the dysregulated nodes highlighted within our network are linked to humoral immunity, complement pathways, extracellular matrix dynamics, and thrombin-mediated coagulation. These perturbations may be provoked through intense immune response during acute infection resulting in endothelial and other organ injury that fails to fully resolve, as suggested through the significantly higher CRP signal at visit 1 and other reports in the literature (12,47,48). Our findings align with similar reports of persistent complement dysregulation and endothelial injury in PASC (11,25,49–51).

If confirmed, these mechanisms could motivate strategies to rebalance coagulation and complement. Hypothesised candidate agents identified via an FDA drug-target overlay, arising from a curated target layer (OpenTargets) in figure 4, may be targets for future controlled studies (no clinical benefit is yet implied). Thrombin has two drugs that have already been, or suggested to be, employed against COVID-19: DABIGATRAN and ARGATROBAN (52,53). ANFIBATIDE acts as a platelet GPIbα antagonist and was previously suggested as a potential therapeutic against the thrombo-cardiovascular disorders associated with acute COVID-19 (54). PEGCETACOPLAN acts as a direct complement C3 inhibitor and is currently prescribed in the treatment of blood clotting diseases (55). Lastly, RIVAROXABAN, an F10 inhibitor, has already been assessed in an open-label, multicentre, randomized, controlled trial to improve clinical outcomes following discharge after COVID-19 related hospitalisation (56). We stress that these therapeutic links are hypotheses generated by a correlation-based network and require direct testing; with that caveat, repurposing these agents, already used or proposed in acute COVID-19, merits investigation for the prevention or treatment of PASC.

These insights emerged only through an integrative, network-based approach: single-omic and genome-wide analyses were largely null, whereas coherent signals appeared at the pathway and node-reorganisation level, reinforcing that PASC reflects multifactorial perturbation of immune–coagulative homeostasis rather than a single molecular change.

## Limitations

Several limitations temper these findings. First, our measurements are predominantly blood-based, which biases detection toward circulating processes and cannot fully represent tissue-level biology in the lung, gut or brain. Second, and most importantly, correlation-based networks, even when consistent across omics, cannot establish whether the complement– coagulation–platelet coupling we observe drives symptoms or is a downstream consequence of another persistent stimulus; the thrombin-centred model we propose is therefore a hypothesis for validation rather than an established mechanism. Third, power is limited: no single-omic feature survived FDR correction, the differential-correlation networks rest on modest group sizes (n = 32 PASC, 45 recovered), and edges estimated at this scale can be unstable, so individual correlations should be read with caution and the rankings regarded as hypothesis-generating. Fourth, recruitment spanned August 2020 to March 2024 and therefore multiple SARS-CoV-2 waves and variants, while evolving vaccination and treatment shifted the case-mix; selection toward patients able to return for follow-up may further limit generalisability. Fifth, with two primary timepoints we cannot establish the temporal ordering needed to claim mechanism. Finally, the multiplex validation is itself constrained by sample size (particularly at convalescence and in the matched subset) and covers only seven markers; its convalescent and interaction trends should be treated as hypothesis-generating, and multiplicity was controlled within, but not across, comparisons.

## Data Availability

All data produced in the present study are available upon reasonable request to the authors.

## Contributors

Conceptualisation: J.C.Y., J.-L.B., L.G., J.d.G., L.B. and L.E.; methodology and investigation: B.W., J.C.Y., J.d.G., J.-L.B., P.D.C., J.-F.C., J.P.D., L.G., V.H., S.J., B.K., S.P.d.R., D.V., L.E. and L.B.; formal analysis and writing – original draft: B.W., L.E. and L.B.; writing – review and editing: all authors; supervision: J.C.Y., J.-L.B., L.G., J.d.G., L.B. and L.E.; funding acquisition: J.C.Y., J.-L.B., L.G., J.d.G., L.B. and L.E. All authors read and approved the final manuscript.

## Data sharing

The multi-omic and clinical datasets generated and analysed in this study are available from the corresponding author on reasonable request, subject to the governance of the HYGIEIA cohort and applicable data-protection regulation; repository and accession identifiers to be added.

## Code availability

The bioinformatic pipelines are provided in Supplementary Information 1; analysis scripts are available from the corresponding author on reasonable request.

## Ethics

The study was conducted in accordance with the Declaration of Helsinki and approved by the Institutional Review Board of Cliniques Universitaires Saint-Luc (protocol 2021/30DEC/543; approved 30 December 2021); it is registered on ClinicalTrials.gov (NCT05557539). All participants provided written informed consent.

## Acknowledgements

The authors thank all patients involved in this study and the clinical research coordinators at Cliniques Universitaires Saint-Luc for their help with recruitment and sample collection.

## References

1. Turner AJ, Hiscox JA, Hooper NM. ACE2: from vasopeptidase to SARS virus receptor. Trends in Pharmacological Sciences. 2004 Jun;25(6):291–4. doi:10.1016/j.tips.2004.04.001

2. Mehta OP, Bhandari P, Raut A, Kacimi SEO, Huy NT. Coronavirus Disease (COVID-19): Comprehensive Review of Clinical Presentation. Front Public Health. 2021 Jan 15;8:582932. doi:10.3389/fpubh.2020.582932

3. Hill EL, Mehta HB, Sharma S, Mane K, Singh SK, Xie C, et al. Risk factors associated with post-acute sequelae of SARS-CoV-2: an N3C and NIH RECOVER study. BMC Public Health. 2023 Oct 25;23(1):2103. doi:10.1186/s12889-023-16916-w

4. Chen C, Haupert SR, Zimmermann L, Shi X, Fritsche LG, Mukherjee B. Global Prevalence of Post-Coronavirus Disease 2019 (COVID-19) Condition or Long COVID: A Meta-Analysis and Systematic Review. J Infect Dis. 2022 Nov 1;226(9):1593–607. doi:10.1093/infdis/jiac136 PubMed PMID: 35429399; PubMed Central PMCID: PMC9047189.

5. Mandel H, Yoo Y, Allen A, Abedian S, Verzani Z, Karlson E, et al. Long COVID incidence in adults and children between 2020 and 2023: a real-world data study from the RECOVER Initiative [Internet]. In Review; 2024 [cited 2025 May 15]. Available from: https://www.researchsquare.com/article/rs-4124710/v1 doi:10.21203/rs.3.rs-4124710/v1

6. Soriano JB, Murthy S, Marshall JC, Relan P, Diaz JV, WHO Clinical Case Definition Working Group on Post-COVID-19 Condition. A clinical case definition of post-COVID-19 condition by a Delphi consensus. Lancet Infect Dis. 2022 Apr;22(4):e102–7. doi:10.1016/S1473-3099(21)00703-9 PubMed PMID: 34951953; PubMed Central PMCID: PMC8691845.

7. Thaweethai T, Jolley SE, Karlson EW, Levitan EB, Levy B, McComsey GA, et al. Development of a Definition of Postacute Sequelae of SARS-CoV-2 Infection. JAMA. 2023 Jun 13;329(22):1934. doi:10.1001/jama.2023.8823

8. Venkatesan P. NICE guideline on long COVID. The Lancet Respiratory Medicine. 2021 Feb;9(2):129. doi:10.1016/S2213-2600(21)00031-X

9. Kenny G, Townsend L, Savinelli S, Mallon PWG. Long COVID: Clinical characteristics, proposed pathogenesis and potential therapeutic targets. Front Mol Biosci. 2023;10:1157651. doi:10.3389/fmolb.2023.1157651 PubMed PMID: 37179568; PubMed Central PMCID: PMC10171433.

10. Raj SR, Arnold AC, Barboi A, Claydon VE, Limberg JK, Lucci VEM, et al. Long-COVID postural tachycardia syndrome: an American Autonomic Society statement. Clin Auton Res. 2021 Jun;31(3):365–8. doi:10.1007/s10286-021-00798-2

11. Mohandas S, Jagannathan P, Henrich TJ, Sherif ZA, Bime C, Quinlan E, et al. Immune mechanisms underlying COVID-19 pathology and post-acute sequelae of SARS-CoV-2 infection (PASC). eLife. 2023 May 26;12:e86014. doi:10.7554/eLife.86014

12. Sherif ZA, Gomez CR, Connors TJ, Henrich TJ, Reeves WB, RECOVER Mechanistic Pathway Task Force. Pathogenic mechanisms of post-acute sequelae of SARS-CoV-2 infection (PASC). eLife. 2023 Mar 22;12:e86002. doi:10.7554/eLife.86002

13. Su Q, Lau RI, Liu Q, Li MKT, Yan Mak JW, Lu W, et al. The gut microbiome associates with phenotypic manifestations of post-acute COVID-19 syndrome. Cell Host & Microbe. 2024 May;32(5):651–660.e4. doi:10.1016/j.chom.2024.04.005

14. Wu X, Xiang M, Jing H, Wang C, Novakovic VA, Shi J. Damage to endothelial barriers and its contribution to long COVID. Angiogenesis. 2024 Feb;27(1):5–22. doi:10.1007/s10456-023-09878-5 PubMed PMID: 37103631; PubMed Central PMCID: PMC10134732.

15. Davis HE, McCorkell L, Vogel JM, Topol EJ. Long COVID: major findings, mechanisms and recommendations. Nat Rev Microbiol. 2023 Mar;21(3):133–46. doi:10.1038/s41579-022-00846-2

16. Peluso MJ, Deeks SG. Mechanisms of long COVID and the path toward therapeutics. Cell. 2024 Oct 3;187(20):5500–29. doi:10.1016/j.cell.2024.07.054 PubMed PMID: 39326415; PubMed Central PMCID: PMC11455603.

17. Chen HJ, Appelman B, Willemen H, Bos A, Prado J, Geyer ChiaraE, et al. Transfer of IgG from Long COVID patients induces symptomology in mice [Internet]. Immunology; 2024 [cited 2025 May 27]. Available from: http://biorxiv.org/lookup/doi/10.1101/2024.05.30.596590 doi:10.1101/2024.05.30.596590

18. Mignolet M, Deroux C, Florkin T, Bielarz V, De Swert K, Halloin N, et al. Pathogenic IgG from long COVID patients with neurological sequelae triggers sensitive but not cognitive impairments upon transfer into mice. Acta Neuropathol. 2026 Apr 29;151(1):50. doi:10.1007/s00401-026-03019-0

19. Santos Guedes De Sa K, Silva J, Bayarri-Olmos R, Brinda R, Alec Rath Constable R, Colom Diaz PA, et al. A causal link between autoantibodies and neurological symptoms in long COVID [Internet]. Allergy and Immunology; 2024 [cited 2025 May 27]. Available from: http://medrxiv.org/lookup/doi/10.1101/2024.06.18.24309100 doi:10.1101/2024.06.18.24309100

20. Saito S, Shahbaz S, Luo X, Osman M, Redmond D, Cohen Tervaert JW, et al. Metabolomic and immune alterations in long COVID patients with chronic fatigue syndrome. Front Immunol. 2024;15:1341843. doi:10.3389/fimmu.2024.1341843 PubMed PMID: 38304426; PubMed Central PMCID: PMC10830702.

21. Su Y, Yuan D, Chen DG, Ng RH, Wang K, Choi J, et al. Multiple early factors anticipate post-acute COVID-19 sequelae. Cell. 2022 Mar 3;185(5):881–895.e20. doi:10.1016/j.cell.2022.01.014 PubMed PMID: 35216672; PubMed Central PMCID: PMC8786632.

22. Wang K, Khoramjoo M, Srinivasan K, Gordon PMK, Mandal R, Jackson D, et al. Sequential multi-omics analysis identifies clinical phenotypes and predictive biomarkers for long COVID. Cell Rep Med. 2023 Nov 21;4(11):101254. doi:10.1016/j.xcrm.2023.101254 PubMed PMID: 37890487; PubMed Central PMCID: PMC10694626.

23. Wei Y, Gu H, Ma J, Mao X, Wang B, Wu W, et al. Proteomic and metabolomic profiling of plasma uncovers immune responses in patients with Long COVID-19. Front Microbiol. 2024;15:1470193. doi:10.3389/fmicb.2024.1470193 PubMed PMID: 39802657; PubMed Central PMCID: PMC11718655.

24. Yang H, Guan L, Xue Y, Li X, Gao L, Zhang Z, et al. Longitudinal multi-omics analysis of convalescent individuals with respiratory sequelae 6-36 months after COVID-19. BMC Med. 2025 Mar 5;23(1):134. doi:10.1186/s12916-025-03971-w PubMed PMID: 40038650; PubMed Central PMCID: PMC11881263.

25. Cervia-Hasler C, Brüningk SC, Hoch T, Fan B, Muzio G, Thompson RC, et al. Persistent complement dysregulation with signs of thromboinflammation in active Long Covid. Science. 2024 Jan 19;383(6680):eadg7942. doi:10.1126/science.adg7942

26. Ward B, Yombi JC, Balligand JL, Cani PD, Collet JF, De Greef J, et al. HYGIEIA: HYpothesizing the Genesis of Infectious Diseases and Epidemics through an Integrated Systems Biology Approach. Viruses. 2022 Jun 23;14(7):1373. doi:10.3390/v14071373

27. Ward B, Pyr Dit Ruys S, Balligand JL, Belkhir L, Cani PD, Collet JF, et al. Deep Plasma Proteomics with Data-Independent Acquisition: Clinical Study Protocol Optimization with a COVID-19 Cohort. J Proteome Res. 2024 Sep 6;23(9):3806–22. doi:10.1021/acs.jproteome.4c00104

28. Michel LYM, Esfahani H, De Mulder D, Verdoy R, Ambroise J, Roelants V, et al. An NRF2/â3-Adrenoreceptor Axis Drives a Sustained Antioxidant and Metabolic Rewiring Through the Pentose-Phosphate Pathway to Alleviate Cardiac Stress. Circulation. 2025 May 6;151(18):1312–28. doi:10.1161/CIRCULATIONAHA.124.067876 PubMed PMID: 40071326; PubMed Central PMCID: PMC12052078.

29. Demichev V, Messner CB, Vernardis SI, Lilley KS, Ralser M. DIA-NN: neural networks and interference correction enable deep proteome coverage in high throughput. Nat Methods. 2020 Jan;17(1):41–4. doi:10.1038/s41592-019-0638-x

30. Kokla M, Virtanen J, Kolehmainen M, Paananen J, Hanhineva K. Random forest-based imputation outperforms other methods for imputing LC-MS metabolomics data: a comparative study. BMC Bioinformatics. 2019 Dec;20(1):492. doi:10.1186/s12859-019-3110-0

31. Bolger AM, Lohse M, Usadel B. Trimmomatic: a flexible trimmer for Illumina sequence data. Bioinformatics. 2014 Aug 1;30(15):2114–20. doi:10.1093/bioinformatics/btu170

32. Kim D, Langmead B, Salzberg SL. HISAT: a fast spliced aligner with low memory requirements. Nat Methods. 2015 Apr;12(4):357–60. doi:10.1038/nmeth.3317 PubMed PMID: 25751142; PubMed Central PMCID: PMC4655817.

33. Liao Y, Smyth GK, Shi W. featureCounts: an efficient general purpose program for assigning sequence reads to genomic features. Bioinformatics. 2014 Apr 1;30(7):923–30. doi:10.1093/bioinformatics/btt656 PubMed PMID: 24227677.

34. Michael Love SA. DESeq2 [Internet]. Bioconductor; 2017 [cited 2025 Nov 4]. Available from: https://bioconductor.org/packages/DESeq2 doi:10.18129/B9.BIOC.DESEQ2

35. Li H. Aligning sequence reads, clone sequences and assembly contigs with BWA-MEM [Internet]. arXiv; 2013 [cited 2025 Nov 4]. Available from: https://arxiv.org/abs/1303.3997 doi:10.48550/ARXIV.1303.3997

36. Li H, Handsaker B, Wysoker A, Fennell T, Ruan J, Homer N, et al. The Sequence Alignment/Map format and SAMtools. Bioinformatics. 2009 Aug 15;25(16):2078–9. doi:10.1093/bioinformatics/btp352

37. DePristo MA, Banks E, Poplin R, Garimella KV, Maguire JR, Hartl C, et al. A framework for variation discovery and genotyping using next-generation DNA sequencing data. Nat Genet. 2011 May;43(5):491–8. doi:10.1038/ng.806

38. Plagnol V, Curtis J, Epstein M, Mok KY, Stebbings E, Grigoriadou S, et al. A robust model for read count data in exome sequencing experiments and implications for copy number variant calling. Bioinformatics. 2012 Nov 1;28(21):2747–54. doi:10.1093/bioinformatics/bts526

39. Cameron DL, Baber J, Shale C, Papenfuss AT, Valle-Inclan JE, Besselink N, et al. GRIDSS, PURPLE, LINX: Unscrambling the tumor genome via integrated analysis of structural variation and copy number [Internet]. Bioinformatics; 2019 [cited 2025 Nov 4]. Available from: http://biorxiv.org/lookup/doi/10.1101/781013 doi:10.1101/781013

40. Cingolani P, Platts A, Wang LL, Coon M, Nguyen T, Wang L, et al. A program for annotating and predicting the effects of single nucleotide polymorphisms, SnpEff: SNPs in the genome of Drosophila melanogaster strain w1118; iso-2; iso-3. Fly (Austin). 2012;6(2):80–92. doi:10.4161/fly.19695 PubMed PMID: 22728672; PubMed Central PMCID: PMC3679285.

41. Rau J, Mitchell J, Fortenberry Y, Church F. Heparin Cofactor II: Discovery, Properties, and Role in Controlling Vascular Homeostasis. Semin Thromb Hemost. 2011 Jun;37(04):339–48. doi:10.1055/s-0031-1276582

42. Pierangeli SS, Girardi G, Vega-Ostertag M, Liu X, Espinola RG, Salmon J. Requirement of activation of complement C3 and C5 for antiphospholipid antibody–mediated thrombophilia. Arthritis & Rheumatism. 2005 Jul;52(7):2120–4. doi:10.1002/art.21157

43. Esmon CT. Regulation of blood coagulation. Biochimica et Biophysica Acta (BBA) - Protein Structure and Molecular Enzymology. 2000 Mar;1477(1–2):349–60. doi:10.1016/S0167-4838(99)00266-6

44. Iglesias MJ, Sanchez-Rivera L, Ibrahim-Kosta M, Naudin C, Munsch G, Goumidi L, et al. Elevated plasma complement factor H related 5 protein is associated with venous thromboembolism. Nat Commun. 2023 Jun 7;14(1):3280. doi:10.1038/s41467-023-38383-y

45. Knopf PM, Rivera DS, Hai S, McMurry J, Martin W, De Groot AS. Novel function of complement C3d as an autologous helper T-cell target. Immunol Cell Biol. 2008 Mar;86(3):221–5. doi:10.1038/sj.icb.7100147

46. Von Andrian UH, Chambers JD, McEvoy LM, Bargatze RF, Arfors KE, Butcher EC. Two-step model of leukocyte-endothelial cell interaction in inflammation: distinct roles for LECAM-1 and the leukocyte beta 2 integrins in vivo. Proc Natl Acad Sci USA. 1991 Sep;88(17):7538–42. doi:10.1073/pnas.88.17.7538

47. Islam MS, Wang Z, Abdel-Mohsen M, Chen X, Montaner LJ. Tissue injury and leukocyte changes in post-acute sequelae of SARS-CoV-2: review of 2833 post-acute patient outcomes per immune dysregulation and microbial translocation in long COVID. Journal of Leukocyte Biology. 2023 Mar 1;113(3):236–54. doi:10.1093/jleuko/qiac001

48. Ward B, Bindels LB, Balligand JL, Bearzatto B, Bommer G, Cani PD, et al. Association of nasopharyngeal *Dolosigranulum pigrum* and *Corynebacterium* species with post-acute sequelae of SARS-CoV-2 in a longitudinal cohort. Claesen J, editor. Microbiol Spectr. 2026 Apr 7;14(4):e02313–25. doi:10.1128/spectrum.02313-25

49. Baillie K, Davies HE, Keat SBK, Ladell K, Miners KL, Jones SA, et al. Complement dysregulation is a prevalent and therapeutically amenable feature of long COVID. Med. 2024 Mar;5(3):239–253.e5. doi:10.1016/j.medj.2024.01.011

50. Bayarri-Olmos R, Bain W, Iwasaki A. The role of complement in long COVID pathogenesis. JCI Insight. 2025 Aug 22;10(16):e194314. doi:10.1172/jci.insight.194314

51. Kell DB, Laubscher GJ, Pretorius E. A central role for amyloid fibrin microclots in long COVID/PASC: origins and therapeutic implications. Biochemical Journal. 2022 Feb 25;479(4):537–59. doi:10.1042/BCJ20220016

52. Cerezo-Manchado JJ, Meca Birlanga O, García de Guadiana Romualdo L, Gil-Ortega I, Martínez Francés A, Iturbe-Hernandez T. Dabigatran in patients with atrial fibrillation after COVID-19 hospitalization: an update of the ANIBAL protocol. Drugs Context. 2022;11:2021-9–4. doi:10.7573/dic.2021-9-4 PubMed PMID: 35145555; PubMed Central PMCID: PMC8798364.

53. El Sobky SA, Fawzy IO, Ahmed MS, Ragheb M, Hamad MHM, Bahaaeldin R, et al. Drug repurposing of argatroban, glimepiride and ranolazine shows anti-SARS-CoV-2 activity via diverse mechanisms. Heliyon. 2025 Feb;11(3):e41894. doi:10.1016/j.heliyon.2025.e41894

54. Chérifi F, Laraba-Djebari F. Bioactive Molecules Derived from Snake Venoms with Therapeutic Potential for the Treatment of Thrombo-Cardiovascular Disorders Associated with COVID-19. Protein J. 2021 Dec;40(6):799–841. doi:10.1007/s10930-021-10019-4 PubMed PMID: 34499333; PubMed Central PMCID: PMC8427918.

55. Griffin M, Kelly R, Brindel I, Maafa L, Trikha R, Muus P, et al. Real-world experience of pegcetacoplan in paroxysmal nocturnal hemoglobinuria. American J Hematol. 2024 May;99(5):816–23. doi:10.1002/ajh.27242

56. Ramacciotti E, Barile Agati L, Calderaro D, Aguiar VCR, Spyropoulos AC, De Oliveira CCC, et al. Rivaroxaban versus no anticoagulation for post-discharge thromboprophylaxis after hospitalisation for COVID-19 (MICHELLE): an open-label, multicentre, randomised, controlled trial. The Lancet. 2022 Jan;399(10319):50–9. doi:10.1016/S0140-6736(21)02392-8

