## Supplement information 1 for "Longitudinal multi-omic network rewiring at the complement– coagulation interface in post-acute sequelae of COVID-19 (PASC)"

### Supplementary information 1: Supplementary methodology

**Patient cohort composition, sample collection, and storage**

Participants for this analysis were selected from the HYGIEIA cohort [1]. The cohort comprised 25 healthy individuals, 24 influenza patients, 50 moderate COVID-19 cases (defined as non-hospitalised, WHO clinical Progression Scale score <4), and 57 severe COVID-19 patients (hospitalized, WHO Clinical Progression Scale score >4) recruited from Cliniques Universitaires Saint-Luc and Grand Hôpital de Charleroi [2]. Enrolment occurred from August 2020 to March 2024. Ethical approval was obtained from the local ethics committee (reference 2021/30DEC/543), and the study protocol is registered on ClinicalTrials.gov (identifier NCT05557539).

Participant samples and medical data were collected at two time points, first at participant inclusion (usually one day after admission for hospitalised patients, or during acute disease for non-hospitalised patients) and again at a second time point during convalescence. 80% of participants completed both visits, resulting in follow-up samples from 24 healthy controls, 16 influenza patients, 46 moderate COVID-19 patients, and 38 severe COVID-19 patients, with an average interval of 81 days between sampling events. At each time point, study nurses collected the following samples/data: one tube whole blood 4 mL EDTA K3E S-Monovette (Sarstedt, Germany); one tube whole blood 4 mL Lithium heparin gel prepared S-Monovette (Sarstedt, Germany); Tempus Blood RNA Tube (Thermo Fisher Scientific, USA); patient signs and symptoms recorded as part of a patient medical questionnaire. Additional information was also collected from medical data bases including full medical histories, pre-existing conditions and comorbidities, medications taken, treatment prescribed before/during hospitalisation, blood-based clinical laboratory data from hospitalisation, physiological vital parameters during hospitalisation/inclusion.

Immediately following sample collection, samples were transported in a cold box to a pre-analytical laboratory for processing. Tempus blood and EDTA blood were aliquoted, flash frozen (liquid nitrogen), and stored at -80°C. Plasma was separated from lithium heparin blood via centrifugation (2600g for 10 minutes at 4°C) (Sigma 3-16KL, Sigma Laboratory Centrifuges, Osterode am Harz, Germany), then aliquoted, flash frozen (liquid nitrogen), and stored at -80°C. Time from sample collection to storage was kept under 2 hours for all sample types.

To avoid unintentional batch effects in the data generation phase, sample batches were randomly created programmatically, ensuring patient sex, cohort group (healthy, flu, mild COVID, severe COVID), sample collection date, and visit number of samples was consistent as regarding the cohort as a whole.

**Clinical and medical parameters**

Clinical data and biological parameters were collected systematically at each study visit by a registered nurse. Additionally, comprehensive clinical histories and patient medical records were obtained via detailed review of digital hospital documentation by clinical researchers. Collected data encompassed patient demographics, clinical severity classifications (WHO Clinical Progression Scale scores), hospitalization details (admission, discharge, ICU stays), medical history, symptomatology (including respiratory, neurological, gastrointestinal, cardiovascular, and general symptoms), comorbidities, medication use, vaccination status, and treatment interventions (oxygen therapy, ventilation support, ECMO, vasopressors, renal replacement, tracheostomy, and antibiotic treatments).

Further, all routine laboratory data from clinical blood and biochemical analyses when performed on the participants during hospitalisation and/or study visits were also recorded. The data encompass coagulation parameters (activated partial thromboplastin time [APTT], prothrombin time [PT], international normalized ratio [INR]), complete blood counts (haemoglobin, haematocrit, erythrocyte parameters, leukocyte differential, platelet counts), inflammatory biomarkers (CRP, procalcitonin, ferritin, fibrinogen, D-dimer), renal and hepatic function tests (urea, creatinine, glomerular filtration rate [eGFR], electrolytes, bilirubin, AST, ALT, alkaline phosphatase), metabolic parameters (glucose, albumin, lipids, lactate dehydrogenase [LDH], creatine kinase [CK]), hormonal assays (TSH, free T4), and infection-specific parameters (SARS-CoV-2 viral load, anti-SARS-CoV-2 antibody levels). Blood gas measurements, including acid-base status and respiratory function (pH, bicarbonate, arterial oxygen and carbon dioxide tensions, oxygen saturation), and specialized hematological and biochemical markers (e.g., troponin-T, NT-proBNP, vitamin D, HbA1c, immunoglobulins, complement levels, and autoantibodies) were also documented.

Wet lab methodology

**DNA extraction and whole exome sequencing**

DNA was extracted from EDTA blood using QIAamp DNA Blood Mini Kit (Qiagen, Germany) in random batches of 16 as previously described. 400 µL EDTA blood and 40 µL QIAGEN protease was combined and homogenised via pipetting. 400 µL buffer AL was then added, pulse-vortexed for 20 seconds, briefly spun-down (3–5 seconds) in a benchtop mini-centrifuge, then incubated for 10 minutes as 56°C. Following, samples were spun down and 400 µL absolute ethanol was added. Samples were then pulse-vortexed for 20 seconds and spun down again briefly. Subsequently, 600 µL of mixture was transferred onto a QIAamp Mini spin column, then centrifuged at 6,000 × g for 1 minute and flow-through was discarded. This step was repeated twice for a total of 1200 µL sample mixture. Columns were then transferred to a new collection tube and 500 µL buffer AW1 was added to each column, followed by centrifugation at 6,000 × g for 1 minute. After discarding flow-through, columns were placed in new collection tubes and washed with 500 µL buffer AW2, followed by centrifugation at 20,000 × g for 3 minutes. Flow-through was discarded, and columns underwent an additional centrifugation at 20,000 × g for 1 minute to remove residual buffer. For DNA elution, columns were transferred to 1.5 mL DNase-free tubes and DNA was eluted by adding 100 µL AE buffer directly onto the membrane, incubating at room temperature (15–25°C) for 5 minutes, then centrifuging at 6,000 × g for 1 minute. Eluted DNA was reapplied to the column, incubated, and centrifuged again to maximize DNA recovery.

Library preparation and sequencing for WES were performed using the Twist Human Core Exome (+RefSeq) system (Twist Bioscience, USA), following the manufacturer's recommended protocols. Briefly, 50 ng total gDNA in 10 µL was added to 40 µL enzymatic fragmentation master mix and placed within a thermo cycler at 32°C for 22 minutes, followed by 65°C for 30 minutes. 5 µL Twist Universal Adapters was then added to each tube, followed by 45 µL of ligation master mix (prepared according to manufacturer) and gently mixed before incubation in thermal cycler at 20°C for 15 minutes. DNA was subsequently purified using 80 µL of the provided DNA magnetic purification beads in two cycles of 200 µL 80% ethanol wash, followed by 5 minutes air-drying. DNA was then eluted from beads using 17 µL buffer EB; 15 µL was then transferred to new tube. Adapter-ligated libraries were PCR amplified using 10 µL Twist Unique Dual Indexed (UDI) Primers and 25 µL

Equinox Library Amp Mix, following recommended conditions (98°C for 45 sec; 7 cycles of 98°C for 15 sec, 60°C for 30 sec, and 72°C for 30 sec; final extension at 72°C for 1 min), then purified with 50 µL of purification beads and eluted in 22 µL Buffer EB as previously described, resulting in 20 µL Amplified Indexed Libraries transferred to a new tube.

Amplified and indexed libraries were quantified and pooled (8-plex) using 187.5ng indexed library (1500 ng total mass per pool). Pools were then vacuum-concentrated with no heat then resuspended in 5 µL blocker solution and 7 µL universal blocker. Hybridisation probe solution was prepared according to manufactures protocol using Core Exome and RefSeq panel. Both probe solution and library pool were then heated to 95°C for 2 minutes and 5 minutes respective, before equilibration to room temperature. Probe solution was then added to the library pool followed by 30 µL Hybridization Enhancer, samples were then hybridized at 70°C for 16 hours. Following hybridization, probe-target complexes were captured on pre-washed streptavidin-coated magnetic beads, gently mixed for 30 minutes at room temperature, then washed once with 200 µL wash buffer 1 and three times with 200 µL wash buffer 2 at 48°C, followed by resuspension in 45 µL water to form bead slurry.

Post-capture amplification was carried out using 22.5 µL Streptavidin Binding Bead Slurry, 25 µL Equinox Library Amplification Mix and 2.5 µL Twist Amplification Primers, with PCR settings according to manufacturer recommendations (98°C for 45 sec; 8 cycles of 98°C for 15 sec, 60°C for 30 sec, 72°C for 30 sec; and a final extension at 72°C for 1 min). The final enriched libraries were purified using 50 µL DNA purification beads, two washes of 200 µL 80% ethanol, and elution in 32 µL buffer EB, of which 30 µL was transferred to a clean tube. Libraries were quality controlled using Picogreen (Thermo Fisher Scientific, USA) and Tapestation D1000 screen tape (Agilent, USA). Sequencing was subsequently carried out on a Novaseq 6000 platform to a target depth of 5Gb, 50x mappable, 151 paired-end reads.

**Whole Transcriptomic Shotgun RNAseq**

Total RNA was isolated from whole-blood samples collected in Tempus RNA tubes using the Tempus™ Spin RNA Isolation Kit (cat.# 4380204, Thermo Fisher Scientific, USA) in batches of 20. Approximately 3 mL of blood was transferred into sterile 15 mL Falcon tubes along with 1 mL of PBS (Ca^2+/Mg^2+-free). The tubes were vortexed for 30 seconds and subsequently centrifuged at 3,000 × g for 30 minutes at 4°C. Following, the supernatant was discarded and tubes inverted 2 minutes to drain residual liquid, after which residual fluid at the rim was gently removed. RNA pellets were resuspended with 400 µL RNA purification resuspension solution, then solution was transferred onto a pre-wetted RNA purification filter column. Samples were incubated on filters for 2 minutes, then centrifuged at 16,000 × g for 30 seconds. The flow-through was discarded, and filters were washed once with 500 µL RNA purification wash solution 1, followed by two washes with 500 µL RNA purification wash solution 2, centrifuging each time at 16,000 × g for 30 seconds. After a final drying centrifugation step at 16,000 × g for 30 seconds, each column was transferred to a new collection tube. RNA was eluted by adding 100 µL Nucleic Acid Purification Elution Solution, incubating for 2 minutes at 70°C, and centrifuging at 16,000 × g for 30 seconds. The elution was repeated by adding the same eluate back onto the filter and centrifuging again at 18,000 × g for 2 minutes. Finally, 90 µL of the eluted RNA was carefully transferred to new RNase-free tubes without disturbing any residual pellet.

Extracted RNA samples underwent further purification using the RNA Clean & Concentrator-5 Kit (cat.# R1013, Zymo Research, USA). Briefly, each sample was treated with 11.25 µL DNA Digestion Buffer and 11.25 µL reconstituted DNase I enzyme, vortexed briefly, and incubated at room temperature for 15 minutes. Post-treatment clean-up involved the addition of 225 µL RNA binding buffer and 450 µL absolute ethanol, followed by loading onto Zymo-spin IC columns in two steps. For each step, 450 µL of the sample was added to the column, centrifuged at 16,000 × g for 30 seconds, and flow-through discarded. Filters were then washed sequentially using 400 µL RNA prep buffer, followed by washes with 700 µL and 400 µL RNA wash buffer, each centrifuged at 16,000 × g for 30 seconds to 1 minute. RNA was finally eluted by adding 25 µL 60°C DNase/RNase-free water to the filter, incubating for 5 minutes at room temperature, and centrifuging at 16,000 × g for 30 seconds each time. Elution step was repeated twice for a total of 50 µL elute.

Library preparation and sequencing for RNA-seq from whole blood were performed using a combination of the Illumina Globin-Zero Gold Kit (Illumina, USA) for rRNA/globin mRNA depletion, followed by the TruSeq Stranded mRNA protocol (Illumina, USA) for library construction. Briefly, 1 µg of DNA-free total RNA in 26 µL RNase-Free Water was combined with 4 µL Globin-Zero Gold Reaction Buffer, and 10 µL Globin-Zero Gold Removal Solution. After thorough mixing, each sample was heated at 68 °C for 10 minutes, briefly cooled to room temperature, then incubated for a further 5 minutes. Following probe hybridisation, 65 µL of thoroughly washed magnetic beads (room-temperature-equilibrated) were added to each reaction. The mixture was then pipette-mixed, rapidly vortexed for 10 seconds, and incubated for 5 minutes at room temperature and an additional 5 minutes at 50 °C. The tubes were then placed on a magnetic stand for 1 minutes, and the supernatant was carefully transferred to new tubes.

Depleted RNA was subsequently purified and concentrated via ethanol precipitation. Sample volumes were adjusted to 180 µL using RNase-Free Water. 18 µL of 3 M sodium acetate and 2 µl of glycogen (10 mg/ml) were then added and the tube gently vortexed, followed by 600 µl of ice-cold 100% ethanol. Tubes were again gently vortexed and placed at -20 °C for 1 hour, then centrifuged at 10,000 x g for 30 minutes, after which supernatant was discarded. The resulting pellet was then washed twice with ice-cold 70% ethanol and then air dried at room temperature for 5 minutes.

For subsequent library preparation, the final pellet was thoroughly dissolved in 18 µL FPF (Fragment, Prime, Finish Mix), followed by fragmentation at 94 °C for 8 minutes in a thermal cycler, cooling on ice, then collection by quick centrifugation. After, add 8 µL of First Strand Synthesis Act D Mix (FSA) containing reverse transcriptase (SuperScript II) to the RNA/FPF mixture and place sample in thermo cycler under recommended conditions (25 °C for 10 minutes, 42 °C for 15 minutes, 70 °C for 15 minutes). After first-strand cDNA synthesis, add 20 µL of Second Strand Marking Master Mix (SMM) to each sample, mix slowly, then incubate at 16 °C for 60 minutes in a thermal cycler. Once the incubation is complete, add 90 µL of resuspend AMPure XP beads and mix thoroughly, then let the mixture incubate at room temperature for 10–15 minutes. After incubation, wash the pellet twice in 200 µL of freshly prepared 80% ethanol, using a magnetic stand and discarding the supernatant each time, then let beads air-dry for about 15 minutes on magnetic stand. Remove samples from the magnetic stand and elute cDNA using 17.5 µL of Resuspension Buffer (RSB), then carefully transfer 15 µL of the supernatant to a fresh tube. Blunt-ended cDNA fragments were adenylated by adding 2.5 µL diluted CTA (A-Tailing Control) and 12.5 µL ATL (A-Tailing Mix) to each sample. The samples were thoroughly mixed and incubated at 37°C for 30 minutes followed by 5 minutes at 70°C. Reactions were rapidly cooled on ice for 1-minute post-incubation. Indexed adapters were ligated to the adenylated cDNA fragments by adding

2.5 µL diluted CTL (Ligation Control), 2.5 µL LIG (Ligation Mix), and 2.5 µL RNA adapters to each sample. Samples were thoroughly mixed, then incubated at 30°C for 10 minutes, after which they were placed on ice and 5 µL STL (Stop Ligation Buffer) was added. Ligated fragments were cleaned using two rounds of AMPure XP bead purification (first with 42 µL, second with 50 µL), in which beads were added to samples, incubated for 15 minutes, washed twice with 200 µL 80% ethanol, and eluted into RSB (first with 52.5 µL, second with 22.5 µL). Finally, 20 µL of RSB was transferred to a fresh PCR plate. Adapter-ligated fragments were enriched using PCR by adding 5 µL PCR Primer Cocktail (PPC) and 25 µL PCR Master Mix (PMM) to each sample, followed by mixing, then thermal cycling with initial denaturation at 98°C for 30 seconds, 15 cycles of amplification (98°C for 10 seconds, 60°C for 30 seconds, 72°C for 30 seconds), and final extension at 72°C for 5 minutes. Post-PCR clean-up involved adding 47.5 µL AMPure XP beads, washing twice with 80% ethanol, and eluting in 32.5 µL RSB, followed by transferring 30 µL elute to a new target sample plate.

Libraries were quality controlled using Picogreen (Thermo Fisher Scientific, USA) and Tapestation D1000 screen tape (Agilent, USA). Final libraries were normalized to a concentration of 10 nM in 10 mM Tris-HCl, pH 8.5 with 0.1% Tween 20. Normalized libraries were subsequently pooled in equal volumes to create a final sequencing pool. Pooled libraries were finally sequenced on a Novaseq 6000 (Illumina, USA) to a target depth of 30M 151 paired-end reads.

**DIA Shotgun Proteomics**

Plasma samples were processed according to our published protocol in batches of 16 [3]. Briefly, 600 µL of High Select Top14 Abundant Protein Depletion Resin (Thermo Fisher Scientific, USA) was equilibrated at room temperature, mixed gently with 22 µL of plasma, and incubated on a rotational mixer (12 rpm) for 10 min at room temperature. The resin-sample mixture was then transferred onto Pierce™ Micro-Spin columns (Thermo Fisher Scientific, USA) and centrifuged at 1000 × g for 2 minutes, with the depleted protein collected as flow-through.

The flow-through then underwent thermal denaturation at 95°C for 5 minutes, then cooled at room temperature. Proteins were reduced by adding 33 µL 50 mM DL-dithiothreitol (5 mM final concentration) and incubating at 56°C for 1 hour with shaking at 1000 rpm.

Alkylation followed by adding 37µl of 500mM chloroacetamide (50 mM final concentration) and incubating for 30 min at room temperature in the dark. Proteins were precipitated using 65.5µl 100% trichloroacetic acid (15% final concentration), incubated on ice for 30 minutes, pelleted at 4000 × g for 7.5 minutes, and the resulting pellet washed three times with 500 µL -20°C acetone. After air-drying for 10 mintues, pellets were resuspended in 75µL 50 mM triethylammonium bicarbonate, sonicated twice at 37kHz pulsed for 2 minutes, and digested overnight with 2.5 µg sequencing grade modified trypsin (V5117, Promega, United States) at 37°C, 750 rpm.

Following digestion, peptides were acidified by adding 8.3 µL of 1% trifluoroacetic acid (final concentration 0.1%) and 80 µg total peptide immediately loaded onto a Pierce High pH Reversed-Phase Peptide Fractionation column (Thermo Fisher Scientific, USA). Columns were centrifuged at 3000 × g for 2 minutes, flow-through discarded, and columns washed once with 300 µL MS-grade water at 3000 × g for 2 minutes. Peptides were sequentially eluted into seven fractions using solutions of increasing acetonitrile concentration (7.5%, 10%, 12.5%, 15%, 17.5%, 20%, and 50%), each containing 0.1% triethylamine. Fractionation was performed by centrifugation at 3000 × g for 2 minutes for each elution step.

Fractions were strategically pooled: fractions 1, 2, and 5 (100 µL each) were combined as pooled fraction 1; fractions 3 and 6 (150 µL each) as pooled fraction 2; and fractions 4 and 7 (150 µL each) as pooled fraction 3. The pooled samples were briefly frozen on dry ice for 5 minutes, then fully evaporated at 4°C using a vacuum concentrator at 4°C. Each dried pooled fraction was resuspended in 20 µL MS resuspension buffer (3.5% acetonitrile, 0.1% trifluoroacetic acid), sonicated three times at 37 kHz pulsed for 2 minutes, briefly vortexed, and spun down prior to LC-MS/MS analysis.

Peptides (1 µg per injection) were loaded onto a reversed-phase pre-column (Acclaim PepMap 100, Thermo Fisher Scientific, USA) and separated using an EasySpray analytical column (Acclaim PepMap RSLC C18, 0.075 × 250 mm, Thermo Fisher Scientific, USA). Separation employed a 120-minute gradient at 300 nL/min on an Ultimate 3000 RSLC nanoHPLC system (Thermo Fisher Scientific, United States): solvent B (0.1% TFA in 80% ACN) was linearly increased from 4% to 32% over 100 minutes, then raised to 50% over 5 minutes, increased further to 90% in the next 5 minutes, and maintained at 95% for the final 9 minutes.

Peptides were detected in the Orbitrap at a resolution of 30,000 (FWHM). MS/MS spectra were acquired using stepped higher-energy collisional dissociation (HCD) at 22%, 26%, and 30% of maximum. A data-independent acquisition (DIA) approach was employed to scan precursor ions across an m/z range of 500–740 with a 4 m/z isolation window at a resolution of 60,000 (FWHM). MS1 spectra were recorded with an automatic gain control (AGC) target of 1.2 × 10^6 ions and a maximum injection time of 55 ms, while MS2 spectra were collected with an AGC target of 1.5 × 10^5 ions and an automatically set maximum injection time, covering an m/z range of 145 to 1450. Each injection had a run time of 120 minutes.

**Shotgun Metabolomics**

Plasma samples were processed in batches of 18. 200 µL plasma was combined with 600 µL -20°C acetone (with internal standards at 50 µM) and vortexed vigorously for 10 seconds. Samples were then sonicated at 37 kHz pulsed for 2 minutes, then centrifuged at 10,000 x g at 4°C for 10 minutes. Carefully, 560 µL of the upper-phase was transferred to a clean tube. For the tube containing the lower-phase and pellet, 600 µL (without internal standards) was added, vortexed vigorously, sonicated at 37 kHz pulsed for 2 minutes, then centrifuged at 10,000 x g for 10 minutes at 4°C. 560 µL of the resulting upper-phase was then combined with the previously extracted upper-phase and vortexed to combine. The upper-phase mixture was then aliquoted into 280 µL samples, and all aliquots were dried down on a heating block at 30°C with nitrogen flush system. Two sample aliquots were then resuspended in 50% acetonitrile/0.1% formic acid, specifically prepared for reverse-phase chromatography. Two additional aliquots were then resuspended in 95% acetonitrile containing 0.1% formic acid and 10 mM ammonium formate, designed for hydrophilic interaction chromatography (HILIC). Samples then underwent centrifugation at 10,000 × g for 5 min at 4°C, after which the supernatants were carefully transferred into analytical vials ready for UPLC-MS injection.

Prepared samples were then analysed in three batches of 100, 97, and 55 using a Synapt-XS quadrupole time-of-flight (Q-ToF) mass spectrometer (Waters, USA), operated in resolution mode, coupled to an Acquity Premier UPLC system (Waters, USA). Chromatographic separations were conducted using an Acquity Premier HSS T3 column (2.1 × 100 mm, 1.8 µm particle size; cat.# 186009468, Waters, USA) for reverse-phase analyses and an Acquity Premier BEH Amide column (2.1 × 100 mm, 1.7 µm particle size; cat.# 186009505, Waters,

USA) for HILIC-based separations. Each chromatographic method employed one aliquot analysed in positive electrospray ionization mode and a second aliquot in negative ionization mode. Data were acquiring MSE mode over an m/z range of 50–1200 with leucine-enkephalin lock-mass.

**Targeted multiplex validation**

As an orthogonal, quantitative validation of the complement–coagulation–inflammatory axis nominated by the discovery omics, seven plasma analytes (C9, C-reactive protein (CRP), coagulation factor VII, alpha-1 antitrypsin (SERPINA1), von Willebrand factor, sICAM-1 and sVCAM-1) were quantified by electrochemiluminescence multiplex immunoassay (Meso Scale Discovery) in the same acute (visit 1) and convalescent (visit 2) plasma samples, each measured in duplicate.

Samples were split across two 96-well plates according to required dilutions. ICAM-1, CRP, VCAM-1, Factor VII, and vWF were placed in spot maps 1, 2, 3, 8, and 10 respectfully and analysed at a sample dilution of 4000-fold. C9 and SERPINA1 were placed in spots 1 and 10 respectfully on a separate plate and analysed at a sample dilution of 200,000-fold.

50µl of each prepared calibrator standard or diluted sample were added to each well. Plates were then sealed and incubated at room temperature (21°C) with shaking at 200RPM for 2 hours. Plates were then washed three times with at least 150 µl per well of 1X MSD Wash Buffer. 50µl of detection antibody solution was then added to each well, plate sealed again, and incubated with identical shaking for 1 hour. Plates were then washed three times with at least 150 µl per well of 1X MSD Wash Buffer. 150µl of MSD GOLD Read Buffer B was then added to each well and the plate was analysed on an MSD instrument.

Bioinformatic pipelines

**Proteomics**

Raw mass spectrometry data (.mzML files) were initially processed using DIA-NN (v1.8.1) [4]. All files related to the same pooled fraction were processed together, thus in three batches, in order to leverage the homogeneous nature of each fraction to optimize peak detection and noise removal for improved identification and quantification. For each fraction, the DIA-NN analysis was performed with an in-silico predicted spectral library generated from the

UniProtKB human reference proteome (accession UP000005640) filtered to include only reviewed (Swiss-Prot) canonical proteins [5]. The analysis was configured for library-free search with FASTA digestion; deep learning–based prediction of spectra, retention times (RTs), and ion mobilities (IMs); and employed customized parameters including a precursor FDR cutoff of 1%, fixed mass ranges for fragment (200–1800 m/z) and precursor ions (300–1800 m/z), N-terminal methionine excision, tryptic digestion with cuts at K* and R*, a maximum of 1 missed cleavage, and variable oxidation of methionine (UniMod:35, +15.994915 Da). Additional parameters were set to enable double-search mode, reanalysis, relaxed protein inference, smart profiling and peak centering.

R (v4.3.1) was used to process the precursor quantification data generated by DIA-NN for the three pooled fractions. Zero values were recoded as missing (NA) and data cleaning was then performed. Here, a logistic regression model was applied to each feature using clinical covariates of interest (cohort disease group, PASC development, and visit) as predictors of feature presence; features with a significant association (adjusted p-value ≤0.01) and less than 75% missing values were classified as missing not at random (MNAR), while non-significant features with less than 30% missing values were classified as missing at random (MAR); all other features were removed. The use of these missing value thresholds ensured that only features with sufficient data coverage were retained for downstream analysis. Next, missing values were imputed using a mixed strategy: random forest imputation for MAR values and quantile regression imputation of left-censored data (QRILC) for MNAR values, ensuring robust recovery of low-abundance signals. The resulting imputed data were merged into a single combined dataset, which was subsequently log-transformed and normalized via variance stabilization normalization (VSN) using QFeatures [6]. Finally, precursor-level intensities were aggregated to the protein level via robust summary method (robust M-estimation) from MsCoreUtils using QFeatures, based on protein group annotation, resulting in the final abundance matrix used for downstream statistical analyses [6,7].

**Metabolomics**

Raw data files generated by the Waters Synapt-XS mass spectrometer were imported into Progenesis QI (V4.2, Nonlinear Dynamics, UK) for pre-processing. Here upon, lock mass correction as performed (based on theoretical m/z of leucine enkephalin) and raw spectra were converted to a 2D ion intensity map. Feature detection was carried out via peak picking, identifying ion features based on accurate mass and retention time (RT). A reference run from a pool representative of all the samples was selected to guide alignment, compensating for RT shifts and ensuring consistent feature matching. Following alignment, the software deconvoluted adducts and grouped isotopes, resulting in a list of retention time–m/z features. This feature list was exported for downstream bioinformatics analyses.

Raw abundance values generated by Progenesis QI were imported into R (v4.3.1). Zero values were recoded as missing (NA), and features were assessed as MNAR or MAR as previously described for proteomics, with the caveat that features present in fewer than 10% of samples within any MS/MS batch were removed to minimise batch effects. Missing values were imputed using random forest imputation for MAR values and QRILC for MNAR values, and imputed data were then log-transformed and normalized using VSN in the same manner as for proteomics. Following this, MS/MS batch correction was performed using the ComBat method from the sva package [8]. Finally, metabolite identifications from Progenesis QI were used to annotate features (if known), and known metabolites were aggregated using a robust summary method as previously described for proteomics, yielding the final abundance matrix for downstream statistical analyses.

**Transcriptomics**

Raw fastq files were initially processed using a standard RNA-seq pipeline, where low-quality bases were trimmed with Trimmomatic (v0.39) and reads were aligned to the GRCh38 human genome using HISAT2 (v2.2.1) [9,10]. Gene-level expression was quantified with featureCounts from Subread (v2.0.3) using the Homo_sapiens.GRCh38.105.chr.gtf annotation [11].

The resulting raw count matrices were loaded into R (v4.3.1), and transformed into a DESeq2 using sample metadata, consisting of biological variables of interest for various design formulas [12]. Variance stabilising transformation (VST) was applied to generate normalized count matrices, which were subsequently used for downstream transcriptomic analyses.

**Genomics**

Raw paired-end fastq files were processed using a complete NGS pipeline to generate variant annotations. Genomic reads were aligned to the human reference genome GRCh38 using BWA-mem (v0.7.17) [13]. Resulting BAM files underwent duplicate removal with SAMTools (v1.12) and base quality recalibration using GATK BaseRecalibrator (v4.2.1) [14,15]. Germline single nucleotide variants (SNVs) and short INDELs were called sample-by-sample using GATK HaplotypeCaller [14]. Structural variations were detected by combining "read depth" (ExomeDepth) with "split read" (GRIDSS-Purple-Linx) [16,17]. The resulting VCF files were normalized with BCFTools (v1.12), filtered according to GATK best practices, and annotated with snpEff (v5.0e) [14,18,19]. Quality control metrics were gathered using FastQC (v0.11.9) and Mosdepth (v0.3.1) [20,21]. Finally, the normalized and filtered VCF files were imported onto the UCLouvain Highlander database, where variants are fully annotated by in-house algorithms (minor allele frequencies, coding impact predictions, splicing effects, conservation scores, and gene function, etc) and public databases (currently Ensembl 100, DBNSFP 4.1 and COSMIC 92) [22].

**Long-COVID multi-omic modelling**

All analyses were conducted in R. For each omic layer, features with near-zero variance were removed using caret::nearZeroVar (default settings) [23]. Proteomic and metabolomic data were analyzed as VSN-normalized intensities; RNA-seq data were analyzed from raw counts using voomWithDreamWeights to obtain observation-level precision weights [24].

For the primary longitudinal analysis, linear mixed models were fit per feature using dream (variancePartition v1.36.3) we tested the PASC status(PASC/recovered) x Visit (1/2) interaction (difference-in-differences: [PASC V2 − PASC V1] − [Recovered V2 − Recovered V1]), with a random intercept for patient ID and covariates (COVID severity (moderate/severe), vaccine status (full vaccination yes/no), sex (male/female), and age (years)) [24]. The pipeline was then re-run for cross-sectional analysis of PASC status at visit 1 and visit 2 independently (not longitudinally) using linear models with empirical-Bayes moderation (limma), adjusting for covariates (Visit 1 analysis: COVID severity (moderate/severe), vaccine status (full vaccination yes/no), sex (male/female), and age (years); Visit 2 analysis: vaccination, sex, and age) [25]. For each fit, per-feature statistics (effect size, standard error, moderated t-statistic, raw p-value) were extracted; multiple testing correction was done via FDR-adjusted p-values. The longitudinal analysis reported statistics for the PASC×Visit interaction contrast; the V1 and V2 analyses reported statistics for the PASC main-effect contrasts.

Multi-omic pathway enrichment was performed separately for V1 and V2 using multiGSEA [26]. Gene-set collections were obtained for KEGG and Reactome with identifiers mapped per layer (transcripts: Ensembl; proteins: UniProt; metabolites: HMDB) [27,28]. For each omic, features were ranked by a signed significance score (direction of the PASC/Visit effect × -log10 p) and tested against KEGG and Reactome gene- and metabolite-sets by per-omic gene-set enrichment (fgsea); per-pathway enrichment p-values were then aggregated across the three omics by the Z-method (Stouffer's) and Benjamini–Hochberg-adjusted across all tested pathways, with pathways at combined FDR < 0.05 considered significant.

**Genomic GWAS and SKAT-O/burden analysis**

Annotated SNP data was exported from the Highlander server and data was filtered according to the following details: all QC metrics passed, only autosomes SNPS, read depth > 10, enforced allelic balance 0.20–0.80, only diploid alleles, and consensus prediction score >= 5 (potentially damaging SNP). Per-sample call rate was then filtered to >= 0.95 and per-SNP call rate was filtered to >= 0.95 (in retained samples). SNPs with maximum allele frequency < 0.05 were removed (cohort MAF) and retained SNPs with Hardy-Weinberg exact test p-value > $1\times10^{-6}$. Monomorphic SNPs were then dropped following filters. Missing genotypes were mean-imputed per SNP (minimal impact at call-rate > 95%). PCA was computed on LD-pruned, position-sorted genotypes ($r^2$ threshold 0.2; window 1000 SNPs; step 200). Two PC outlier were identified via mahalanobis distance but kept as results were stable with/without inclusion (high Spearman ρ of −log10 p). PC1 was found to account for most of the captured variation, including ethnicity. For the final GWAS model, we tested the association between SNP dosage (0-2, additive) and PASC status (PASC vs recovered) using logistic regression, adjusting for age (years), sex (male/female), and ancestry (PC1). Calibration was assessed with genomic inflation factor λGC and QQ plot.

Following, we tested per-gene burden/variance effects using a gene-level aggregation analysis (SKAT-O from SKAT package) [29]. SNPs were mapped to their associated gene symbols to form per-gene sets (minimum two SNPs per gene). A logistic SKAT null model was built using the same GWAS covariates (Age, Sex, PC1) and case status coded 0/1 (recovered/PASC). SKAT-O was then run using predictive damaging scores (as calculated within Highlander), coinciding with the estimated damage of the SNP (scores were winsorized at the 99th percentile, log1p-transformed, rescaled to [0,1], and floored at $1e^{-3}$).

**Network Construction and Multi-Omic Integration**

Proteomic, metabolomic, and transcriptomic abundance matrices were analysed separately in the following pipeline. For each omic, differential abundance correlation testing between recovered and PASC was performed using Differential Gene Correlation Analysis (DGCA) (ddcorALL) [30]. Pearson correlation was used, permutation testing was not used for edge inference. A-priori filtered removed edges with weak correlation or insignificant in both groups (<0.3 absolute correlation, adjusted p-value > 0.01); multiple-testing correction (FDR) was then performed on this reduced set. The resulting z-score difference is used as edge weights to represent the strength of association change.

For each node, we then summarize the magnitude and confidence of its interaction changes between PASC and recovered by aggregating the edge weights of all incident edges within the current network. For each node *n* with incident edges *E*(n) and weights $W_e$, the raw rewiring score *R*(*n*) is the Euclidean norm:

$$R(n) = \sqrt{\sum_{e \in E(n)} w_e^2}$$

Because node degree differs markedly across omic layers (e.g., RNA >> protein), the raw rewiring score is degree dependent. To overcome this, a degree-matched z-score (DMZ) was then employed to normalise the raw rewiring score within specific degree-matched bins *B(i)* (defined through $\log_2$(degree +1)). Within each bin, raw rewiring scores were z-scored, reflecting how extreme a node's rewiring is relative to degree-matched peers.

$$Z_i = \frac{R(n) - \mu B(i)}{\delta B(i)}$$

Following the creation of the differential correlation networks for each omic, the separate networks were then combined, and prior knowledge networks were incorporated. Metabolite–enzyme edges were incorporated by merging highly predicted protein-metabolite interactions from Recon3D with manually curated KEGG pathway maps [28,31]. Protein–protein interactions were extracted from STRING using all detected proteins from our experiments (~1600) as seed nodes and expanding to all first neighbours (physical subnetworks only, very high confidence (> 900)) [32]. Experimentally validated transcription-factor gene regulations were sourced from the manually curated TRRUST v2 database [33].

miRNA-mediated post-transcriptional interactions were imported from DIANA-TarBase v8, composed of experimentally supported miRNA targets on protein-coding transcripts with a microT score > 0.7 [34]. Drug-target information was included using the known drug database from OpenTargets, a database of any approved or clinical candidate drug, and its known targets [35]. Finally, a layer incorporating biology central dogma was included, whereby each protein-coding transcript was additionally linked to its translation product, allowing logical traversal between RNA and protein layers.

**Targeted multiplex validation bioinformatics**

PASC and recovered patients were compared per analyte at each visit using the Mann–Whitney U test (with a log-scale Welch t-test as sensitivity analysis), and effects were re-estimated in covariate-adjusted linear (log10 concentration) and logistic (PASC status) models using the discovery covariate set (acute severity at visit 1, vaccination, sex and age). The acute response was additionally stratified by severity, and matched patients were analysed with a Visit × PASC linear mixed model mirroring the dream framework.

### References


1. Ward B, Yombi JC, Balligand JL, et al. HYGIEIA: HYpothesizing the Genesis of Infectious Diseases and Epidemics through an Integrated Systems Biology Approach. *Viruses*. 2022;14(7):1373. doi:10.3390/v14071373
2. Soriano JB, Murthy S, Marshall JC, Relan P, Diaz JV, WHO Clinical Case Definition Working Group on Post-COVID-19 Condition. A clinical case definition of post-COVID-19 condition by a Delphi consensus. *Lancet Infect Dis*. 2022;22(4):e102-e107. doi:10.1016/S1473-3099(21)00703-9
3. Ward B, Pyr Dit Ruys S, Balligand JL, et al. Deep Plasma Proteomics with Data-Independent Acquisition: Clinical Study Protocol Optimization with a COVID-19 Cohort. *J Proteome Res*. 2024;23(9):3806-3822. doi:10.1021/acs.jproteome.4c00104
4. Demichev V, Messner CB, Vernardis SI, Lilley KS, Ralser M. DIA-NN: neural networks and interference correction enable deep proteome coverage in high throughput. *Nat Methods*. 2020;17(1):41-44. doi:10.1038/s41592-019-0638-x
5. The UniProt Consortium, Bateman A, Martin MJ, et al. UniProt: the Universal Protein Knowledgebase in 2023. *Nucleic Acids Research*. 2023;51(D1):D523-D531. doi:10.1093/nar/gkac1052
6. Laurent Gatto, Christophe Vanderaa. QFeatures. doi:10.18129/B9.BIOC.QFEATURES
7. RforMassSpectrometry Package Maintainer, Laurent Gatto, Johannes Rainer, Sebastian Gibb, Adriaan Sticker, Sigurdur Smarason, Thomas Naake. MsCoreUtils. doi:10.18129/B9.BIOC.MSCOREUTILS
8. Wang M, Huang J, Liu Y, Ma L, Potash JB, Han S. COMBAT: A Combined Association Test for Genes Using Summary Statistics. *Genetics*. 2017;207(3):883-891. doi:10.1534/genetics.117.300257
9. Bolger AM, Lohse M, Usadel B. Trimmomatic: a flexible trimmer for Illumina sequence data. *Bioinformatics*. 2014;30(15):2114-2120. doi:10.1093/bioinformatics/btu170
10. Kim D, Langmead B, Salzberg SL. HISAT: a fast spliced aligner with low memory requirements. *Nat Methods*. 2015;12(4):357-360. doi:10.1038/nmeth.3317
11. Liao Y, Smyth GK, Shi W. featureCounts: an efficient general purpose program for assigning sequence reads to genomic features. *Bioinformatics*. 2014;30(7):923-930. doi:10.1093/bioinformatics/btt656
12. Michael Love SA. DESeq2. Published online 2017. doi:10.18129/B9.BIOC.DESEQ2
13. Li H. Aligning sequence reads, clone sequences and assembly contigs with BWA-MEM. *arXiv*. Preprint posted online 2013. doi:10.48550/ARXIV.1303.3997
14. DePristo MA, Banks E, Poplin R, et al. A framework for variation discovery and genotyping using next-generation DNA sequencing data. *Nat Genet*. 2011;43(5):491-498. doi:10.1038/ng.806
15. Li H, Handsaker B, Wysoker A, et al. The Sequence Alignment/Map format and SAMtools. *Bioinformatics*. 2009;25(16):2078-2079. doi:10.1093/bioinformatics/btp352
16. Plagnol V, Curtis J, Epstein M, et al. A robust model for read count data in exome sequencing experiments and implications for copy number variant calling. *Bioinformatics*. 2012;28(21):2747-2754. doi:10.1093/bioinformatics/bts526
17. Cameron DL, Baber J, Shale C, et al. GRIDSS, PURPLE, LINX: Unscrambling the tumor genome via integrated analysis of structural variation and copy number. *Bioinformatics*. Preprint posted online September 25, 2019. doi:10.1101/781013
18. Cingolani P, Platts A, Wang LL, et al. A program for annotating and predicting the effects of single nucleotide polymorphisms, SnpEff: SNPs in the genome of Drosophila melanogaster strain w1118; iso-2; iso-3. *Fly (Austin)*. 2012;6(2):80-92. doi:10.4161/fly.19695
19. Li H. A statistical framework for SNP calling, mutation discovery, association mapping and population genetical parameter estimation from sequencing data. *Bioinformatics*. 2011;27(21):2987-2993. doi:10.1093/bioinformatics/btr509
20. Pedersen BS, Quinlan AR. Mosdepth: quick coverage calculation for genomes and exomes. Hancock J, ed. *Bioinformatics*. 2018;34(5):867-868. doi:10.1093/bioinformatics/btx699
21. FastQC. Published online June 2015. https://qubeshub.org/resources/fastqc
22. Helaers R, Vikkula M. Highlander: variant filtering made easy. *F1000 Research Limited*. Preprint posted online 2018. doi:10.7490/F1000RESEARCH.1116108.1
23. Kuhn M. Building Predictive Models in *R* Using the **caret** Package. *J Stat Soft*. 2008;28(5). doi:10.18637/jss.v028.i05
24. Hoffman GE, Schadt EE. variancePartition: interpreting drivers of variation in complex gene expression studies. *BMC Bioinformatics*. 2016;17(1):483. doi:10.1186/s12859-016-1323-z
25. Gordon Smyth [Cre A. limma. Published online 2017. doi:10.18129/B9.BIOC.LIMMA
26. Sebastian Canzler, Jörg Hackermüller. multiGSEA. doi:10.18129/B9.BIOC.MULTIGSEA
27. Milacic M, Beavers D, Conley P, et al. The Reactome Pathway Knowledgebase 2024. *Nucleic Acids Research*. 2024;52(D1):D672-D678. doi:10.1093/nar/gkad1025
28. Kanehisa M. KEGG: Kyoto Encyclopedia of Genes and Genomes. *Nucleic Acids Research*. 2000;28(1):27-30. doi:10.1093/nar/28.1.27
29. Wu MC, Lee S, Cai T, Li Y, Boehnke M, Lin X. Rare-variant association testing for sequencing data with the sequence kernel association test. *Am J Hum Genet*. 2011;89(1):82-93. doi:10.1016/j.ajhg.2011.05.029

30. McKenzie AT, Katsyv I, Song WM, Wang M, Zhang B. DGCA: A comprehensive R package for Differential Gene Correlation Analysis. *BMC Syst Biol*. 2016;10(1):106. doi:10.1186/s12918-016-0349-1
31. Brunk E, Sahoo S, Zielinski DC, et al. Recon3D enables a three-dimensional view of gene variation in human metabolism. *Nat Biotechnol*. 2018;36(3):272-281. doi:10.1038/nbt.4072
32. Szklarczyk D, Gable AL, Lyon D, et al. STRING v11: protein-protein association networks with increased coverage, supporting functional discovery in genome-wide experimental datasets. *Nucleic Acids Res*. 2019;47(D1):D607-D613. doi:10.1093/nar/gky1131
33. Han H, Cho JW, Lee S, et al. TRRUST v2: an expanded reference database of human and mouse transcriptional regulatory interactions. *Nucleic Acids Research*. 2018;46(D1):D380-D386. doi:10.1093/nar/gkx1013
34. Karagkouni D, Paraskevopoulou MD, Chatzopoulos S, et al. DIANA-TarBase v8: a decade-long collection of experimentally supported miRNA–gene interactions. *Nucleic Acids Research*. 2018;46(D1):D239-D245. doi:10.1093/nar/gkx1141
35. Buniello A, Suveges D, Cruz-Castillo C, et al. Open Targets Platform: facilitating therapeutic hypotheses building in drug discovery. *Nucleic Acids Research*. 2025;53(D1):D1467-D1475. doi:10.1093/nar/gkae1128
